# Multimodal blood based profiling reveals insights into mechanisms of immunotherapy resistance

**DOI:** 10.1101/2025.04.20.25325955

**Authors:** Samuel J. Wright, Izabella Zamora, Milan Parikh, Deepika Yeramosu, Marijana Rucevic, Moshe Sade-Feldman, Thomas J. LaSalle, Emily M. Blaum, Baolin Liu, Lynn Bi, Sarah Kang, Steven M. Blum, Ngan Nguyen, Jamey Guess, Amrita Kar, Alexis Schneider, David Lieb, Elliot Woods, William Michaud, Aleigha R. Lawless, Tatyana Sharova, Sonia Cohen, Gyulnara Kasumova, Michelle S. Kim, Alexandra-Chloé Villani, Ryan J. Park, Russell W. Jenkins, Samuel J. Klempner, Ryan J. Sullivan, Keith T. Flaherty, Nir Hacohen, Arnav Mehta, Genevieve M. Boland

**Author notes:** Co-corresponding authors: **Nir Hacohen, PhD**, Massachusetts General Hospital Cancer Center 55 Fruit St, Boston, MA 02114, **Genevieve Boland, MD, PhD**, Massachusetts General Hospital Cancer Center 55 Fruit St, Boston, MA 02114, **Arnav Mehta, MD, PhD**, Massachusetts General Hospital Cancer Center 55 Fruit St, Boston, MA 02114.

## Abstract

Many cancer patients treated with immune checkpoint blockade (ICB) do not have durable treatment responses. Circulating biomarkers have the potential to identify patients with primary resistance or early progression on therapy to alter treatment course and avoid unnecessary toxicity. Unbiased multimodal proteomic profiling in blood has been underexplored due to the previously limited scalability of multiplexing technologies or cohorts lacking time-series sampling. To address this, we performed plasma proteomic profiling of >2,900 proteins and high-dimensional mass cytometry of peripheral blood lymphocytes across serial time points in 250 metastatic melanoma patients on ICB treatment. We further obtained 92 patient-matched tumor samples, which were processed for single-cell and/or bulk RNA sequencing. Proteins upregulated post-ICB were associated with inflammatory pathways involving the activation of effector immune functions. Expression of genes corresponding to these proteins was higher in immune cells involved in recruitment and tumor reactivity. Expression of genes corresponding to plasma proteins more abundant in non-responders was highest in suppressive myeloid subsets and malignant cells. We further posit the involvement of these non-responder genes in immunosuppressive and pro-tumor interactions, which we confirmed using publicly available spatial transcriptomic data. We also found that epithelial-specific proteins in the circulation of responders post-ICB correlate with patient toxicity and likely originate from healthy tissues. Together, these data represent one of the deepest peripheral biomarker studies using paired blood and tumor samples in melanoma patients treated with ICB, and begin to elucidate the complex interplay between tumors and the systemic immune response within the host.

## Introduction

Immune checkpoint blockade (ICB) has revolutionized the treatment of many immunogenic tumors^1,2^, and advanced melanoma has had encouraging results with >50% first-line response rates with combination ICB^3^. Early identification of non-responders is essential to mitigate unnecessary treatment toxicities and for alternative treatment approaches. However, predictive biomarkers for response are incompletely characterized and may be unique to patient subsets^4,5^. Several approaches have leveraged clinical variables and tissue biomarkers including tumor mutational burden (TMB), PD-L1 abundance, fraction of copy number alterations, HLA-I loss of heterozygosity (LOH), microsatellite status, interferon signatures, and immune composition from bulk RNA-sequencing^4–10^. While these have proven fruitful, the performance has limited clinical utility due to suboptimal predictive performance, the need for adequate tissue, and a failure to capture a global measure of host response.

Circulating biomarkers provide easy access for serial monitoring and can provide insight into the mechanisms of response to ICB^11–14^. In particular, peripheral blood analysis has revealed unique insight into the functional states and clonality of T cells after ICB^15–17^, but studies assessing systemic effects of ICB on host immune and tumor interactions are limited^18^. Importantly, changes in the plasma proteome in cancer patients treated with ICB are poorly understood^19^. Two recent studies demonstrated a role for IL-8 as a potential predictor of non-response to ICB^20,21^ and paired single-cell analysis from tumor and blood tissue suggests that myeloid cells may be a primary source for circulating IL-8^21^. IL-8 blockade has also been added to ipilimumab plus nivolumab treatment in advanced melanoma, though there was no statistically significant improvement in either overall response rate or progression free survival over ipilimumab plus nivolumab alone^22^. However, the use of plasma proteomics to reveal biological insight into immune responses has been limited by small sample sizes or targeted approaches focused on either individual or small subsets of cytokines^19,23^.

To address these issues, we performed plasma proteomics analysis on 465 samples from a cohort of 250 melanoma patients undergoing immune checkpoint blockade to identify biomarkers of response and resistance that reveal insights into the tumor microenvironment. Using linear mixed effects models, we found proteins associated with response, resistance, and treatment time point, and leveraged patient-matched single-cell RNA sequencing data to infer their relationship to tumor microenvironmental processes. We identified several pro-tumor processes involving fibroblasts, immune cells and melanoma cells related to circulating proteins that were more abundant in non-responders. We also identified toxicity-related processes related to circulating proteins that were more abundant in responders in response to immunotherapy. Together, these results shed light on the capability of circulating proteins to monitor response in melanoma patients undergoing immune checkpoint blockade therapy.

## Results

### Plasma proteomic profiling performed on metastatic melanoma cohort undergoing ICB

We obtained 465 plasma samples from a cohort of 250 melanoma patients with advanced unresectable or metastatic disease undergoing immune checkpoint blockade (ICB) therapy. Of these patients, 148 received single-agent ICB, while 82 received combination ICB therapy (**Fig. S1, Table S1**). Samples were obtained at baseline (pre-ICB) and at a 6-week follow-up (post-ICB), and were subsequently subject to antibody-based proteomic profiling of 2943 proteins (Olink Proteomics, proximity extension assay, **Table S2**). In addition, peripheral blood immune cells in circulation from paired plasma samples were characterized using mass cytometry (Teiko Bio, **Table S3**) to quantify relative proportions and abundance of different immune cell subsets. Patient-matched tumor samples were also obtained for 78 patients, and subject to either (or both of) bulk RNA-sequencing or single-cell RNA sequencing (**Fig. 1a**).

**Figure 1.**
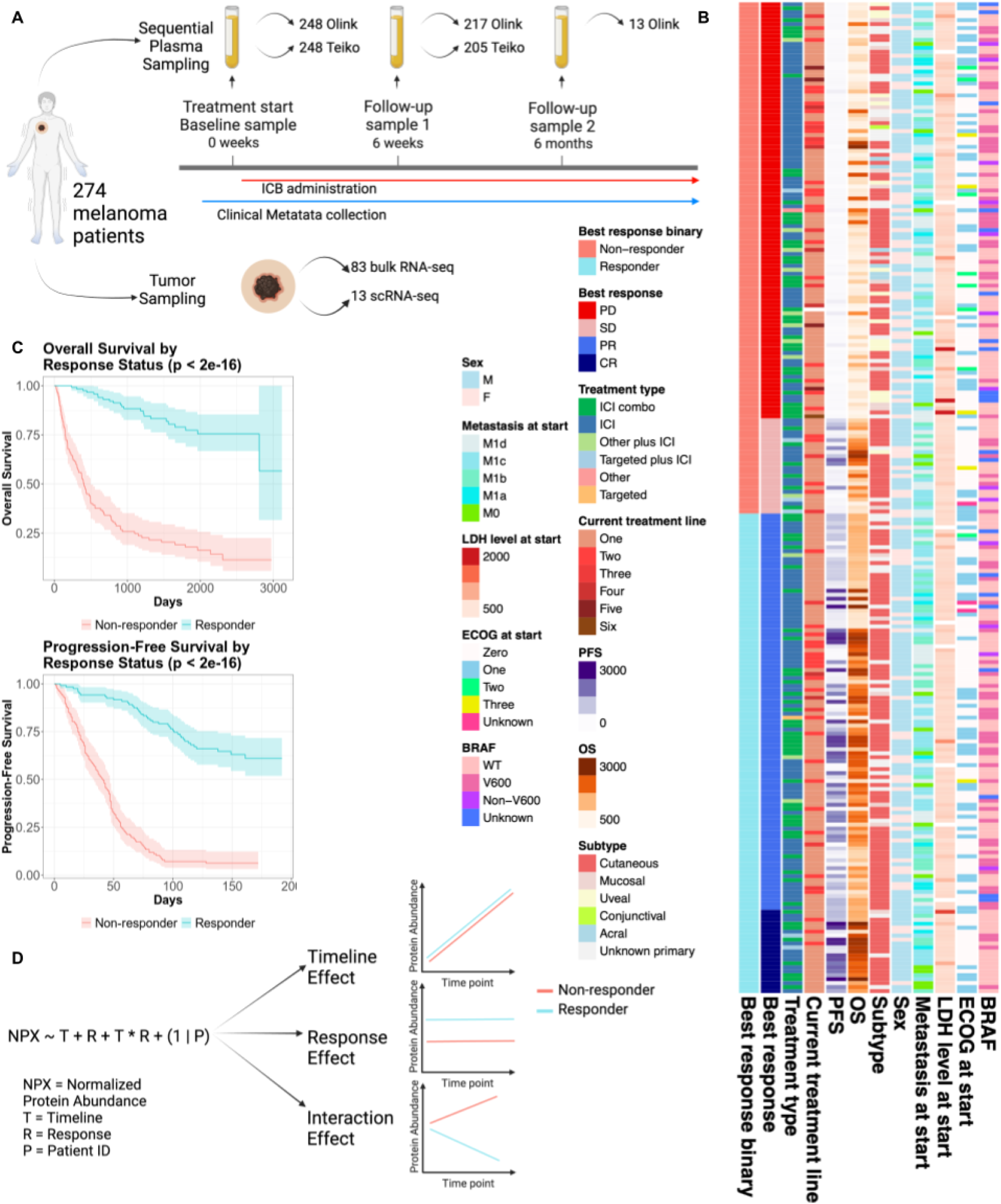
Overview of melanoma proteomics ICB cohort and discovery of time point and response-associated proteins. **a,** Diagram of sample collection from baseline and follow-up treatment time points in addition to parallel tumor sampling, with data modalities listed next to corresponding sampling sites/time points (tumor samples not matched with plasma samples). **b,** Clinical summary of Olink plasma proteomics cohort, including all metadata available at the time of analysis. **c,** Kaplan-Meier curve showing progression-free survival (left) and overall survival (right) of responders (RECIST PR and CR) and non-responders (RECIST SD and PD). Log Rank test. Overall survival, p < 2e-16. Progression free survival, p < 2e-16. **d,** Outline of analysis for discovery of time point and response effects among Olink plasma proteins.

We first analyzed our plasma proteomic dataset to discover proteins associated with time point (post-vs. pre-ICB) effects and response (response vs. non-response) effects. Clinical metadata were collected (**Fig. 1b, Table S1**), and responder or non-responder designation for each patient was made based on their best overall response (BoR) at 6 months following treatment using RECIST criteria. Patients who achieved a BoR of progressive disease (PD) or stable disease (SD) were considered non-responders (NR) (n=129 patients) and patients who achieved a best response of partial response (PR) or complete response (CR) were considered responders (R) (n=121 patients) for subsequent analyses. To quantify survival differences between these two groups, we fit Cox proportional hazards models for the patients’ progression-free survival (PFS) and overall survival (OS) binarized response as a predictor. We found that there were significant differences in the PFS and OS between the groups (**Fig. 1c**).

We performed statistical tests on the clinical metadata to determine whether there was a significant correlation between any of the clinical covariates and response (**Table 1**). There was no significant difference in any of the covariates (except for tumor subtype) when testing either the baseline or post-ICB samples. When considering the distribution of tumor subtypes, the vast majority of responders had cutaneous melanoma, while non-responders were enriched for uveal, mucosal and acral melanoma; as expected, these non-cutaneous subtypes that have poorer rates of response and are genomically distinct^4,24^.

**Table 1.** Clinical covariates’ association with binarized response. Statistical association of clinical covariates with response status (Wilcoxon rank sum test or Fisher’s exact test). Clinical variable with categories listed in the leftmost column. To the right are quantifications of numbers of responder and non-responder samples in each category of the variable (or median with standard deviation) at baseline and post-ICB. P-values for baseline and post-ICB samples are in the rightmost columns.

| Variable | BL R | BL NR | 6W R | 6W NR | p-value (BL) | p-value (6W) |
| --- | --- | --- | --- | --- | --- | --- |
| **Treatment type** |  |  |  |  | 0.150 | 0.200 |
| ICI | 72 | 76 | 63 | 65 |  |  |
| ICI combo | 43 | 38 | 39 | 31 |  |  |
| other + ICI | 2 | 9 | 3 | 9 |  |  |
| targeted + ICI | 2 | 4 | 2 | 4 |  |  |
| targeted | 1 | 0 | 0 | 0 |  |  |
| other | 0 | 1 | 0 | 1 |  |  |
| **Current Tx line** |  |  |  |  | 0.272 | 0.353 |
| 1 | 92 | 88 | 82 | 74 |  |  |
| 2 | 22 | 23 | 20 | 23 |  |  |
| 3 | 4 | 7 | 3 | 6 |  |  |
| 4 | 2 | 4 | 2 | 3 |  |  |
| 5 | 0 | 4 | 0 | 3 |  |  |
| 6 | 0 | 1 | 0 | 1 |  |  |
| **Sex** |  |  |  |  | 0.133 | 0.114 |
| F | 32 | 46 | 30 | 43 |  |  |
| M | 88 | 82 | 77 | 67 |  |  |
| **Age at start** |  |  |  |  | 0.660 | 0.499 |
| Median | 66 ±14.33 | 65 ±14.83 | 64 ±14.40 | 65 ±14.77 |  |  |
| **LDH at start** |  |  |  |  | 0.593 | 0.858 |
| Median | 61.5 ±37.19 | 64 ±44.58 | 59.5 ±33.76 | 57 ±39.60 |  |  |
| **BRAF** |  |  |  |  | 0.409 | 0.552 |
| V600 | 39 | 36 | 35 | 33 |  |  |
| WT | 69 | 70 | 60 | 58 |  |  |
| non-V600 | 4 | 9 | 4 | 9 |  |  |
| unknown | 8 | 13 | 8 | 10 |  |  |
| **Subtype** |  |  |  |  | 1.283e-6 | 2.797e-5 |
| cutaneous | 92 | 73 | 81 | 66 |  |  |
| mucosal | 5 | 13 | 6 | 10 |  |  |
| unknown primary | 23 | 19 | 20 | 14 |  |  |
| acral | 0 | 6 | 0 | 6 |  |  |
| conjunctival | 0 | 1 | 0 | 1 |  |  |
| uveal | 0 | 16 | 0 | 13 |  |  |
| **ECOG at start** |  |  |  |  | 0.095 | 0.356 |
| 0 | 72 | 77 | 66 | 67 |  |  |
| 1 | 44 | 39 | 37 | 34 |  |  |
| 2 | 1 | 7 | 1 | 5 |  |  |
| 3 | 1 | 4 | 1 | 3 |  |  |
| **Metastasis at start** |  |  |  |  | 0.151 | 0.387 |
| M0 | 10 | 8 | 9 | 8 |  |  |
| M1a | 10 | 11 | 9 | 10 |  |  |
| M1b | 30 | 17 | 25 | 15 |  |  |
| M1c | 42 | 53 | 39 | 43 |  |  |
| M1d | 28 | 39 | 25 | 34 |  |  |

When visualizing the distribution of mean normalized protein abundance across the 2943 proteins, it was expected that the means be normally distributed and that the magnitude of the proteomic mean would correspond to a coefficient of variation (CoV) higher in magnitude, which was reflected in the data (**Fig. S2a-b**).

### Identification of treatment time point- and response-associated proteins

To find differentially abundant proteins over time and between response groups in our plasma proteomic dataset, we fit linear mixed effects (LME) models to the abundance values of each of the 2943 proteins in the panel using binarized response, time point, and an interaction term as covariates, with patient ID as a random effect to account for patient-level differences in protein abundance. We evaluated the significance of each covariate in each model and performed multiple hypothesis correction to obtain a finalized list of proteins with any combination of significant time point, response and interaction effects in both positive and negative directions. (**Fig. 1d**).

Initial analysis addressed how treatment affects protein abundance and which proteins are associated with response or non-response to therapy. Proteins were grouped according to their significance for time, response, interaction, or some combination of effects; two proteins had an interaction effect (abundances in responders and non-responders moving in different directions between baseline and post-ICB). 320 and 8 proteins were more abundant post-ICB and pre-ICB, respectively. 27 and 58 proteins were more abundant in responders and non-responders, respectively. (**Fig. 2a-b, Table S4**).

**Figure 2.**
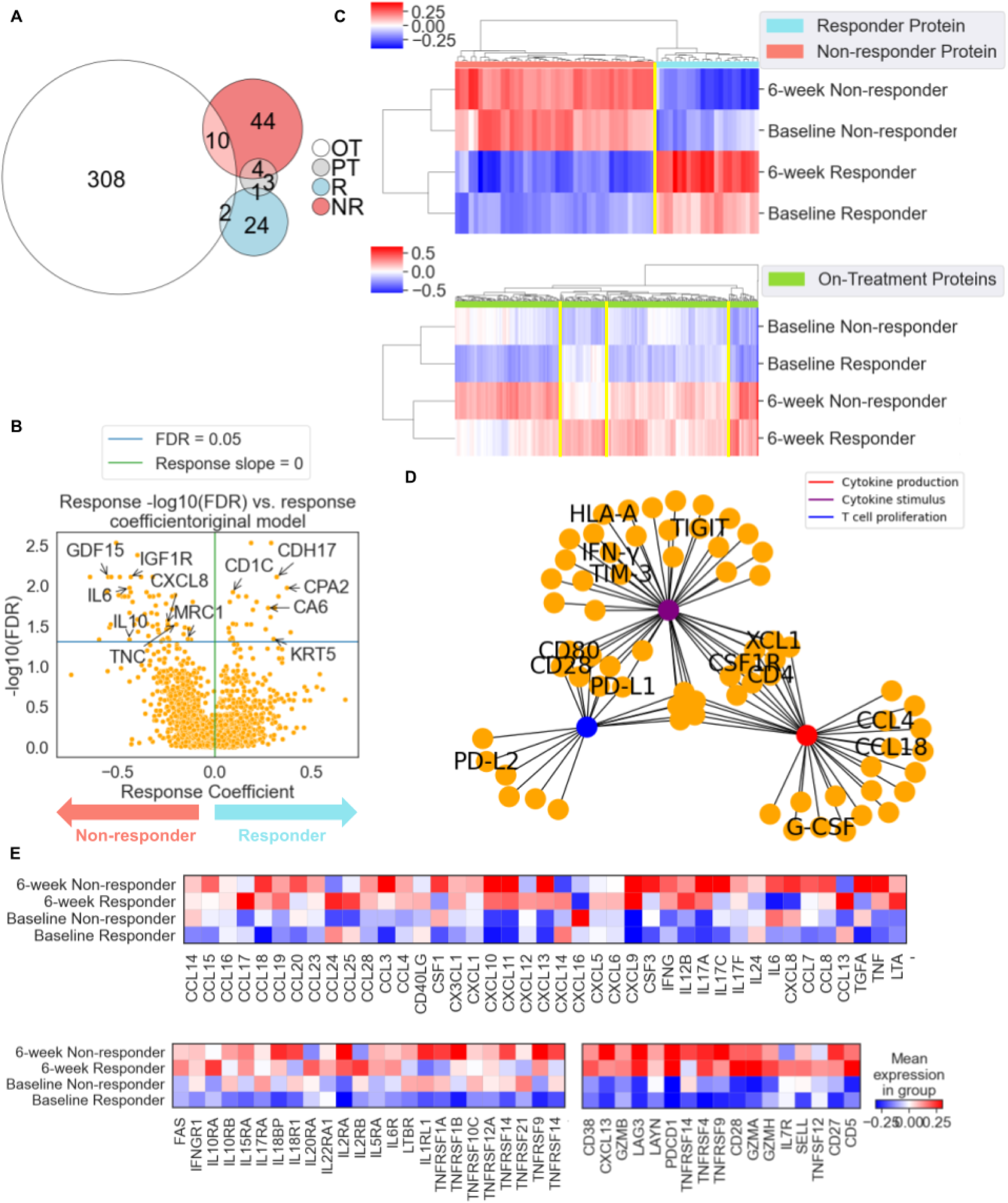
Analysis of circulating proteins discovered through linear mixed effects models points to activation of immune processes induced by ICB. **a.** Venn diagram showing number of proteins with significant time point or response effects. Overlaps show proteins with more than one significant effect. **b,** Volcano plot showing linear mixed effects model response coefficients in x-axis and −log10(FDR) in y-axis. **c,** Heatmaps showing distribution of protein abundances of response effect proteins (top) and post-ICB proteins (bottom). Average group protein abundances are z-scored along the columns. **d**, Network plot representation of gene ontology (GO) enrichment results on-treatment proteins using the GO Biological Processes 2023 library, with orange nodes representing timeline proteins belonging to certain biological processes, represented by non-orange nodes they are connected to. **e,** Heatmaps showing average abundance of immune-related proteins across patient groups split by time point and response. Cytokines and other immune receptors (top), cytokines, chemokines and other secreted immune ligands (bottom left), and T cell-related proteins (bottom right). Average group protein abundances are z-scored along the columns.

We refit the linear mixed regression models using subtype as an additional covariate and noted that most time point-associated proteins remained the same suggesting that this variable did not account for the time point-associated proteomic changes. For response-associated proteins, 8 of the 27 remained significant (CDH17, ITGB6, BMP4, CPA2, MYOC, ITGAV, LEFTY2, CD1C, HMCN2), while for non-response-associated proteins, only 2 of 58 remained significant (STC1, MIA) suggesting that there may be more distinct mechanisms of nonresponse across histologies. The 137 proteins that were associated with subtypes included IGF1R, GDF15, TYRO3, BCAN and PODXL2 (**Fig. S3, Table S5-6**).

We also refit the linear mixed regression models with only cutaneous samples, and interestingly no proteins had significant response effects (**Table S7**). We do note, however, that the proteins which had significant response effects in the original models have highly correlated slopes with the same proteins in the cutaneous-only models (the cutaneous-only model response slopes tended to be lower in magnitude) (**Fig. S4a**), indicating that there is likely some association between the abundances of these proteins and response that fails to reach the level of significance. When comparing the skewness of these protein distributions in responder samples in the overall setting to the cutaneous-only setting, we note that these are similar across response proteins (**Fig. S4b**). However, we noticed that the skewness in the overall setting was higher than the skewness in the cutaneous-only (**Fig. S4c**). In other words, an arm of the protein distribution of non-responder samples that differentiates it from responder samples is lost when non-cutaneous samples are removed from the models.

There is a precedent for including stable disease with responders, so the linear mixed regression models were refitted using SD patients as responders instead of non-responders, and again no proteins had significant response effects (**Table S8**). Again we see a high correlation between the response effect slopes of the response-associated proteins in the original model and the response effect slopes of these proteins in the SD responder models (**Fig. S5a**) Among the responder samples, the skewness in the distribution of these proteins is essentially even (**Fig. S5b**), but in non-responder samples the skewness in the distribution of these proteins is in general slightly higher in the original models, compared to the SD responder models (**Fig. S5c**). This again brings up the possibility that the protein abundance distribution in non-responders is losing an arm that differentiates it from responder samples when SD samples are moved from the non-responder to the responder group.

We found that the abundance of responder and non-responder proteins clustered based on patient response, irrespective of sample time point (**Fig. 2c**). When clustering the abundances of on-treatment proteins in samples based on treatment time point and response status, we found that while all proteins were more highly abundant on-treatment than pre-treatment, the clustering formed four modules of proteins (**Fig. 2c**). This suggests that there may be additional proteins trending towards higher abundance in certain response groups that did not reach the level of significance for response in the linear mixed effects models.

Gene ontology (GO) enrichment analysis using the GO Biological Processes 2023 database revealed that the proteins that increased in abundance after treatment were involved in immune-related biological processes, such as cytokine production, cytokine stimulus and T cell proliferation (**Fig. 2d, Fig. S6**). This notion is further supported by the increased abundance of canonical immune-related proteins in plasma following ICB such as cytokine and chemokines, their receptors, and T cell-related proteins (**Fig. 2e**). Additionally, we noticed that response-dependent differences in abundance between these proteins appeared to be exacerbated after treatment (**Table S4**).

We identified modules of co-abundant proteins (using non-negative matrix factorization) on the post-ICB proteins related to inflammation, such as chemokines/T cell exhaustion, cytotoxicity/proliferation and macrophage processes (**Table S9**). This further emphasizes the relevance of the post-ICB proteins discovered in upregulated immune processes in response to immunotherapy.

Among proteins more abundant in non-responders at both time points are IL6 and IL8 (CXCL8) which have been reported before to be preferentially abundant in circulation in non-responders in melanoma, renal cell carcinoma and urothelial carcinoma^20,21,25,26^. Chemokines classically related to IFNγ response such as CXCL9, CXCL10, CXCL11 and IFNγ are also more abundant post-ICB^27^. CXCL16, which was one of the three pre-ICB proteins, was recently shown to play important roles in tumor immunity and resistance^28,29^. Strikingly, the canonical immune-related proteins that were most uniformly increased in abundance post-ICB in both responders and non-responders were T cell-related proteins such as GZMA, GZMB, CXCL13 and PDCD1. This led us to hypothesize that there was an increased presence of T cells in circulation following ICB regardless of response status.

To leverage our relative circulating immune cell proportions data, we correlated modules of proteins from our linear mixed effects models (response proteins, non-response proteins, and four hierarchically clustered post-ICB protein modules shown in **Fig. 2c**) using cosine similarity with the relative immune cell proportions from our PBMC data. We noticed specific patterns of correlation between the protein modules and immune proportions, such as CD8 T effector memory cells and CD8 TEMRA cells correlating with the response proteins, and Tregs correlating with the non-response proteins (**Fig. S7a**).

We additionally fit linear mixed effects models on the relative immune proportions from our PBMC data using the same strategy as with the Olink NPX values, in which we used patient response, time point and an interaction term to predict each relative immune proportion (**Fig. S7b, Table S10**). While no proportions had terms that reached the level of significance following multiple hypothesis correction, some proteins had uncorrected p-values below 0.05 and thus showed a trend toward significance. Interestingly, response was significantly associated with naive CD8 T cells, indicating that the abundance of this subset in circulation may have some bearing on favorable melanoma ICB outcomes.

Ki67 levels in PD-1+CD8+ cells have been reported to be associated with response to anti-PD1 therapy^16^, and as such we examined our immune cell proportion data to determine whether there was a correlation between Ki67 levels in PD-1+ CD8+ cells in post-ICB samples. Normalizing cell subpopulation fractions by tumor burden has been used as an approach to find correlations with response^16^. We elected to perform this normalization using LDH, which is a more clinically actionable measurement of tumor bulk. Indeed, we noticed that there was a significant correlation between the baseline patient composite score of these circulating melanoma proteins and baseline LDH level in these patients, and the melanoma protein score had a less constrained range than the LDH level, signifying that this may be a more nuanced reflection of tumor bulk (**Fig. S7c**). This approach recapitulated PD1+ CD8+ TEMRA cells having significantly different proportions of Ki67+ cells between responders and non-responders (**Fig. S7d)**, and additionally showed a strong trend of responders having overall PD1+CD8+ cells having higher proportions of Ki67+ cells compared to non-responders (**Fig. S7e**).

To understand whether non-response proteins had associations with other non-cancer diseases (notably other immune-related pathologies), we employed disease clustering annotations provided by Deng et al. 2024^30^. We searched for genes associated with various clusters that were also in the non-responder protein list, and found that for several clusters most non-response proteins were associated with these (**Fig. S8a-b**). We found that GDF15, which has been shown to interfere with antitumor immune response^31^, tended to be the non-responder protein that was most strongly associated with a given cluster, along with resistance-associated cytokines and chemokines such as IL6, CXCL8 and IL10 appearing high in association with many clusters as well (**Table S11**). In examining which diseases were most associated with top overlapping clusters, such as Cluster 34, we noticed that these clusters tend to be associated with immune-related diseases such as sepsis, influenza and gout (**Table S12**).

### Multimodal integration of plasma proteomic and tumor scRNAseq data to identify potential sources of plasma proteins in the tumor

Next, we sought to nominate the source of circulating proteins within the TME and better infer the role of circulating factors in cell-cell interactions within the TME. We hypothesized that a subset of proteins originating from the TME may be detected in circulation. To address this, we generated single-cell RNA-seq data from patient-matched tumor samples (**Fig. 3a, Fig. S9**), to determine which cell types expressed the genes corresponding to proteins with time point, response and interaction effects.

**Figure 3.**
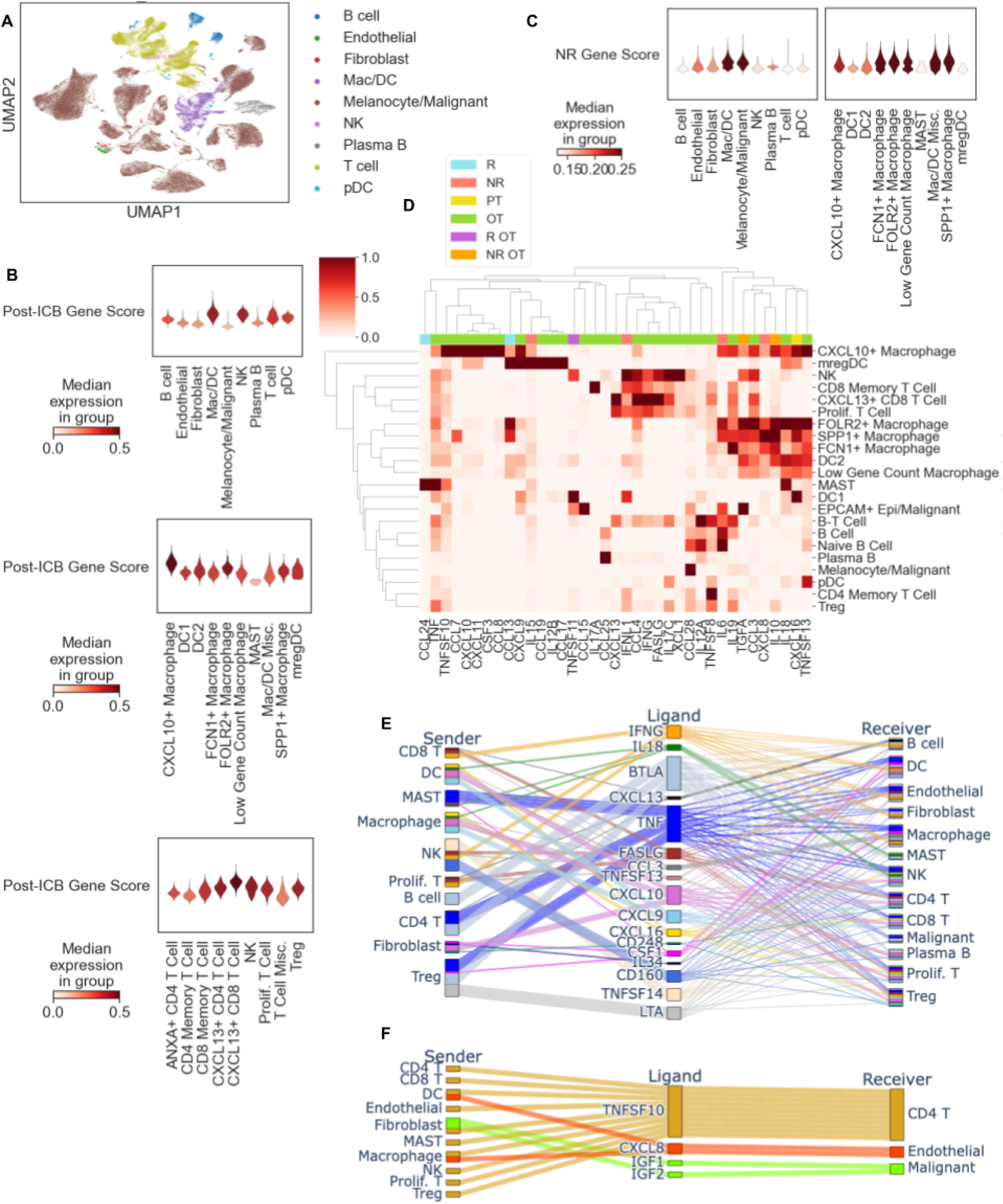
scRNA-seq analysis of time point and response-associated proteins suggests role of immune cells in response and resistance mechanisms within the tumor. **a,** UMAP visualization of single-cell RNA-seq data set, colored by major cell types. **b,** Violin plot visualization of combined score of genes corresponding to upregulated post-ICB proteins in different major cell types (left), macrophage/DC subtypes (middle) and T cell subtypes(right). **c,** Violin plot visualization of combined score of genes corresponding to upregulated non-responder against responder proteins in different major cell types (left) and macrophage and DC subsets (right). **d,** Heatmap visualization of expression of cytokines, chemokines and other secreted immune-related genes across single-cell subtypes. **e,** Flow diagram showing potential interactions involving cytokines and chemokines among post-ICB proteins based on minimum expression of ligands and receptors in sender and receiver cells, respectively. **f,** Flow diagram showing potential interactions involving cytokines and chemokines among non-responder proteins based on minimum expression of ligands and receptors in sender and receiver cells, respectively.

In line with our GO enrichment analysis, the aggregated expression of transcripts from scRNAseq data (using Scanpy score_genes) corresponding to the post-ICB proteins (whether associated with response or non-response) was highest in immune cell subsets such as macrophages and DCs, NK cells and T cells. Within specific immune subsets, expression was highest in CXCL10+ macrophages among macrophages/DCs and CXCL13+ T cells among T cells (**Fig. 3b**). CXCL10+ macrophages are known to attract T cells and other immune cells to the TME via the CXCL10/CXCR3 axis to carry out effector mechanisms^27,32^. CXCL13+ marks tumor-reactive T cells^33^. Altogether, this suggests that these subsets may be contributors to proteins observed in plasma after ICB treatment.

Genes corresponding to non-responder proteins scored highest in macrophages/DCs and malignant cells/melanocytes (**Fig. 3c**). Interestingly, among macrophages and DCs, these non-response-associated genes scored highest in SPP1+ macrophages, which is known as an immunosuppressive macrophage subset^34^.

We then analyzed individual genes corresponding to proteins (with significant response or time point effects), first focusing on cytokines/chemokines (**Fig. 3d, Fig. S10a**). For post-ICB proteins: CXCL9 was found in mregDCs and CXCL10+ macrophages while CXCL10 was expressed highly only in CXCL10+ macrophages; IFNγ and CXCL13 were expressed highly across a number of T cells, especially CXCL13+ CD8 T cells. For non-responder proteins, CXCL8 and IL10 were highly expressed in SPP1+ macrophages, reinforcing the potential importance of these cells in supporting an unfavorable microenvironment that drives resistance. Finally, CXCL16 which is known to interact with CXCR6 on T cells^28,29^ to mediate processes related to tumor immunity, was highly expressed in macrophages and DCs, which is in line with myeloid cells secreting this chemokine^28,29^.

To deepen our understanding of interactions that may involve these cytokines and chemokines in the TME, we performed a ligand-receptor analysis in which we thresholded the expression of cytokines, chemokines and their receptors across cell types to infer cell type pairs that may be participating in a known ligand-receptor interactions. In using this approach to analyze post-ICB proteins that included cytokine/chemokines or cytokine/chemokine receptors, we found several inflammatory signaling axes, including IFNγ signaling (from T and NK cells to myeloid, B and stromal cells)^27^ and CXCL9/10 signaling (from myeloid and stromal cells to T cells)^27^, among many others (**Fig. 3e**). This further supports the hypothesis that these proteins are more abundant in circulation post-ICB as a result of immune recruitment to the tumor, as suggested by the increased abundance of immune recruitment-related proteins (**Fig. 2d**).

Using the same thresholding approach on interactions involving cytokines/chemokines and their receptors among non-responder proteins, we found a subset of interactions that may relate to resistance, including IGF1/2-IGF1R signaling between fibroblasts and tumor cells (**Fig. 3f**). Interestingly, when this approach was adapted to only threshold expression on the receptors participating in these pathways, we noted that there is evidence of several additional cytokine/chemokine receptors despite there being low expression of the cytokine/chemokine source in the TME. These receptors may participate in resistance-associated signaling, such as the IL6^25^ and IL10^35^ pathways (**Fig. S10b**). Although the expression in the TME is low, the preferential abundance of IL6 and IL10 in the plasma of non-responders suggests that there may be unique sensitivity to these ligands regardless of their origin.

### Cell-cell interaction inference highlights the importance of fibroblasts in promoting a pro-tumor microenvironment that can confer resistance

To learn more about potential interactions occurring in the TME that are upregulated in non-responders, we employed CellPhoneDB^36^ to infer interactions involving all proteins on the list of non-responder proteins (**Fig. 4a, Fig. S11-12, Table S13**). CellPhoneDB found evidence of interactions that were identified using the previous threshold ligand-receptor analysis (**Fig. 3e-f**). Additionally, CellPhoneDB inferred other interactions not involving cytokines or chemokines, including the TNC-integrin complex axis and the GAS6-TYRO3 axis. These axes, addition to the IGF1/2-IGF1R axis, involve signaling from fibroblasts to malignant cells, and can contribute to cancer cell growth and metastasis^37–40^. In addition, the PTPRC from T cells signaling to MRC1 on myeloid cells is related to myeloid cell plasticity and may promote the transition of macrophages to more immunosuppressive states^41,42^.

**Figure 4.**
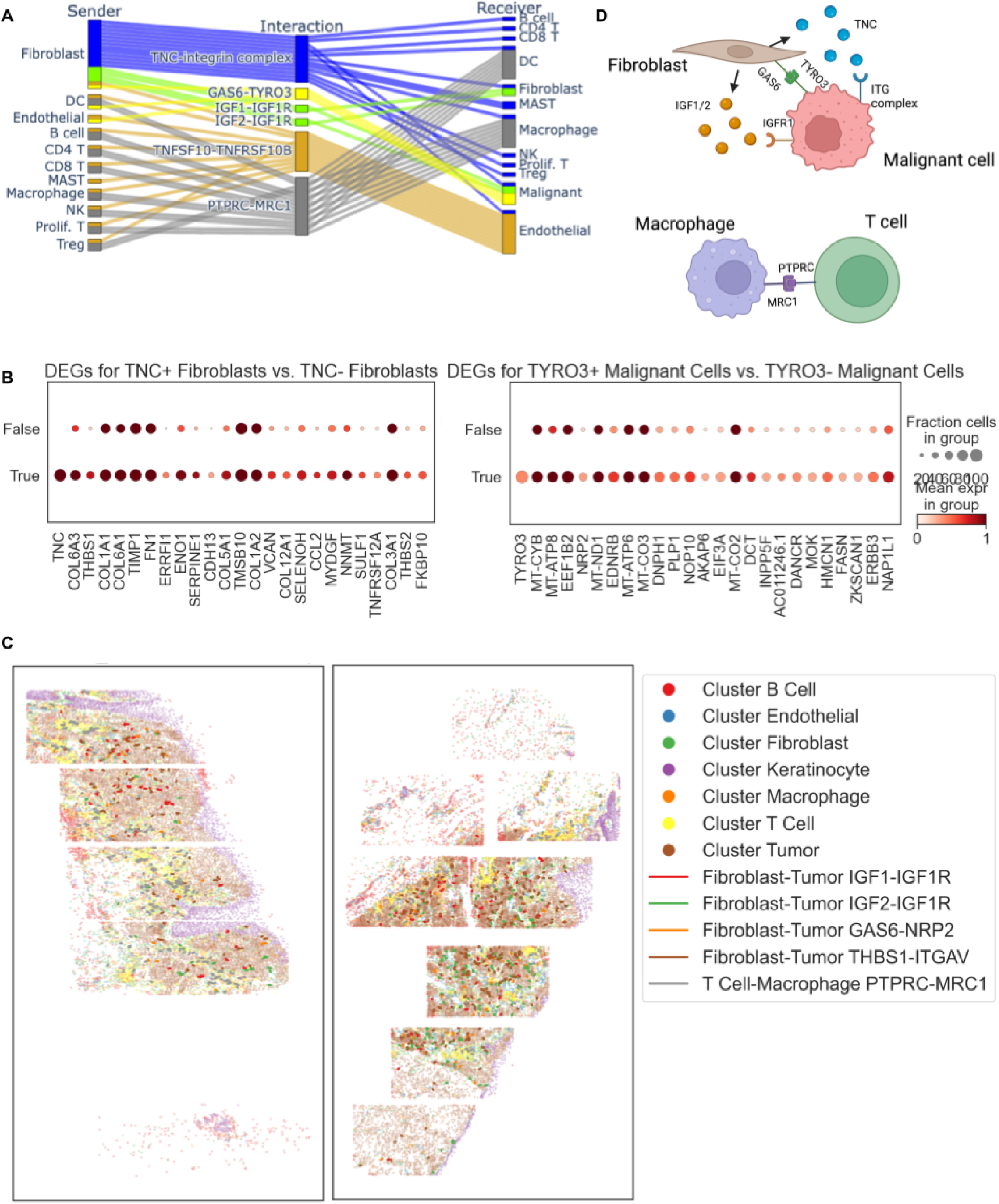
Cell-cell interaction inference highlights importance of fibroblasts in promoting pro-tumor microenvironment that can confer resistance. **a.** Flow diagram showing relevant CellPhoneDB results involving non-responder proteins. **b,** Dotplots showing top differentially expressed genes (DEGs) between TYRO3+ and TYRO3-malignant cells (top) and TNC+ and TNC-fibroblasts (bottom). **c,** Spatial transcriptomic maps showing likely interactions of relevance occurring within melanoma tumors. **d,** Schematic showing potential pro-tumor microenvironment interactions occurring among fibroblasts, immune and malignant cells.

To test whether the cells expressing these ligand-receptor pairs are likely to be adjacent in space in the melanoma TME, we obtained publicly available melanoma CosMx data^43^. For genes that were not available in the CosMx panel (TYRO3 and TNC), we used the scRNA-seq data to identify co-expressed proxy genes (in TYRO3+ malignant cells and TNC+ fibroblasts) (**Fig. 4b)**. We counted the number of adjacent cells with expression of specific ligand-receptor pairs, and performed permutation tests to estimate the significance of each interaction over a null distribution, and found support for several interactions (IGF1/2-IGF1R, GAS6-TYRO3 (NRP2), TNC (THBS1)-ITGAV, and PTPRC-MRC1) in the melanoma TME (**Fig. 4c)**. Pairs of cells expressing PTPRC-MRC1 are co-localized in immune rich regions. For the fibroblast-malignant interactions (TNC-ITGAV, IGF1/2-IGF1R and GAS6-TYRO3), cells co-localized throughout the tumor, with fibroblasts frequently interspersed among malignant cells.

Altogether, these interactions seem to point to potentially pro-tumor (IGF1/2-IGF1R, TNC-ITGAV) and immunosuppressive (GAS6-TYRO3, PTPRC-MRC1) interactions, with a subset of proteins upregulated in the plasma of non-responders (IGF1R, MRC1, TYRO3, TNC), and with interactions among T cells, myeloid cells, stromal cells and malignant cells (**Fig. 4d**).

### Post-ICB responder proteins are associated with clinical toxicity in non-tumor tissues

To learn more about distinguishing features of ICB response in circulation, we turned to post-ICB proteins that appeared to be enriched in responders (**Fig. 2c**). As a group however, the aggregate abundance of these proteins in circulation was higher in responders compared to non-responders at both baseline and on-treatment (**Fig. 5a**). For further analysis, we included the one protein, CPA2, significantly more abundant in both responders ***and*** post-ICB samples. Dubbing these post-ICB responder proteins, we examined their expression in the scRNA-seq data (**Fig. 5b**).

**Figure 5.**
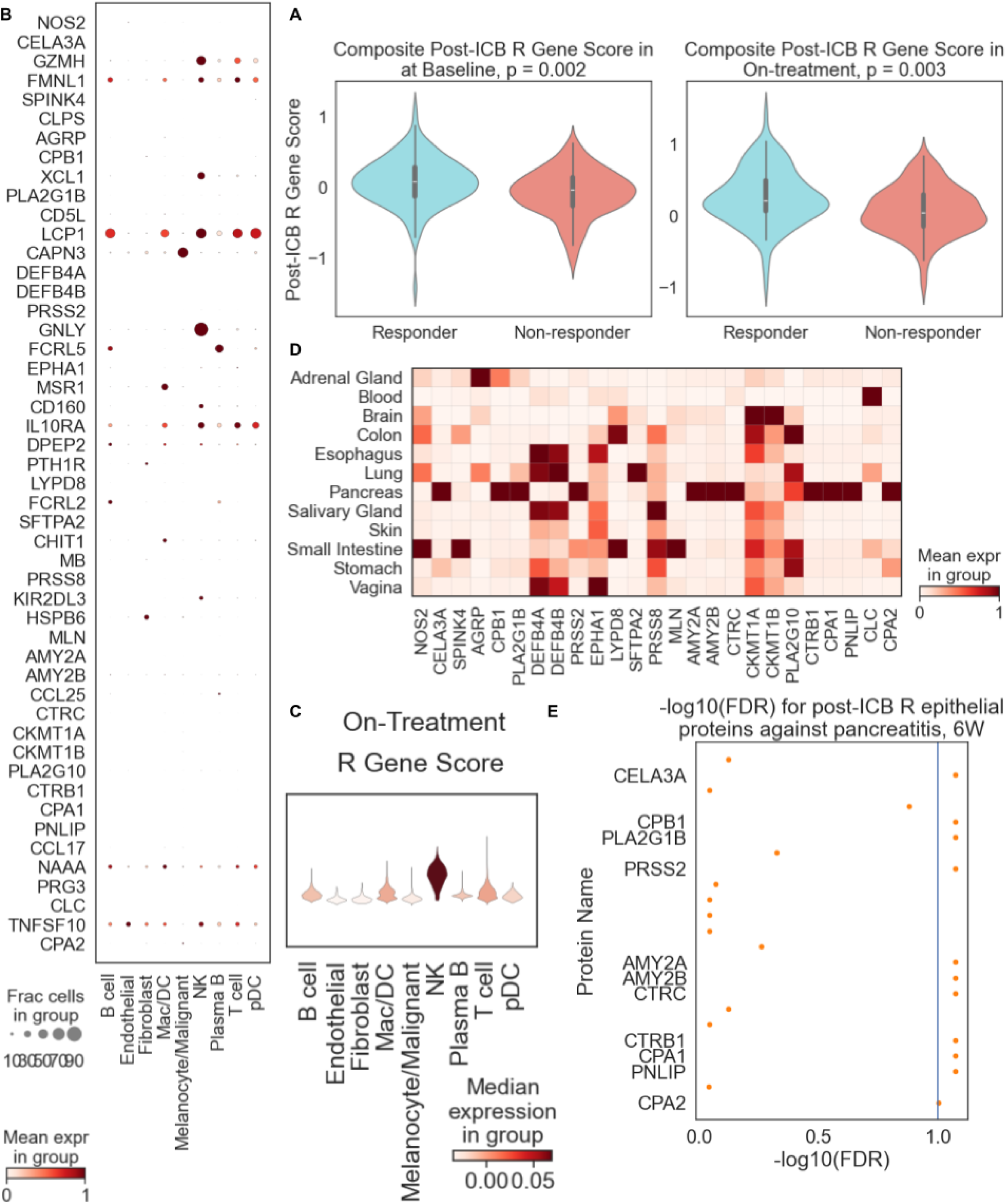
Post-ICB responder proteins are associated with clinical toxicity in non-tumor tissues. **a,** Violin plots showing composite score of post-ICB responder module proteins in responders and non-responders at baseline (left) and on-treatment (right). **b,** Dotplot showing single-cell expression of post-ICB responder proteins across major cell types. **c,** Violin plot visualization of scores for post-ICB responder proteins across major cell types. **d,** Dotplot showing gene expression of post-ICB responder proteins in different tissues across GTEX samples. **e,** Negative log_10_(p-value) of association between post-ICB protein abundance in Olink data and pancreatitis status (Wilcoxon log-rank sum). Only significant proteins are labeled (FDR < 0.1).

We noticed a number of cytotoxic and NK-related genes such as GNLY, XCL1 and GZMH. Indeed, when we scored these genes across cells the highest-scoring cells were NK cells (**Fig. 5c**). This suggests that post-ICB R proteins are involved in NK and cytotoxic processes in the tumor that may promote superior tumor killing and in turn support better clinical response.

We were interested in several other post-ICB responder proteins corresponding to genes that seemed to have little or no expression in the TME. Upon closer examination, we noticed that many of these absent genes, such as CPA1/2, PRSS2 and PLA2G10 are generally expressed on epithelial cells based on GTEX bulk RNA expression^41^, especially pancreas, colon, esophagus and small intestine (**Fig. 5d**). We wondered why proteins from non-malignant tissues were preferentially appearing in responder circulation following ICB. In considering that cytotoxic proteins were also upregulated in the post-ICB responder subset, we hypothesized that cytotoxic processes may be upregulated globally in responders, which in addition to improved cytotoxic response in the tumor may also result in damage in other tissues associated with toxicity, thereby releasing epithelial proteins into circulation.

In an effort to establish a connection between these circulating epithelial proteins and toxicity, we obtained pancreatitis annotations from the patient cohort and correlated the abundance of these proteins with pancreatitis. Of note, the 11 proteins that were most enriched in the pancreas in the GTEx data were significantly associated with pancreatitis (**Fig. 5e**). This suggests that these proteins may originate from tissues outside the tumor which are subject to toxicity due to responder-favored autoimmune cytotoxicity post-ICB. This further demonstrates the capacity of circulating proteins to reveal biological processes originating from not only the tumor but also from normal host tissues in ways that are relevant to response and resistance.

## Discussion

We leverage an integrated analysis of plasma proteomics across serial time points, relative immune PBMC proportions and tissue scRNAseq in melanoma patients treated with immune checkpoint blockade to gain insight into the biology of immunotherapy response and non-response, offering a multi-modal longitudinal dataset of circulating and tumoral transcriptomic and proteomic data. Our analysis reveals sets of proteins that are significantly associated with treatment time points, patient response status, or both. Post-ICB-associated proteins relate ontologically to immune recruitment and response, and we see that groups of immune-related proteins such as cytokines and chemokines, their receptors and T cell markers are all preferentially abundant in post-ICB samples. Our data reveals that canonical cytokines and chemokines that are implicated in favorable responses within the tumor microenvironment show consistent patterns of abundance in the periphery, either reflecting escape from the tumor into circulation or a parallel globalized host response for tumor cells not interacting with the tumor. Indeed, immune recruitment- and activation-related cytokines^27^ such as CXCL9, CXCL10 and CXCL11 along with IFNγ have distinct cellular sources in the tumor microenvironment based on our scRNA-seq, highlighting these immune cells within the TME as sources of these circulating cytokines post-ICB. The increased abundance of these inflammatory signals following ICB, relating to immune cell recruitment and effector mechanisms in the TME, points to a common immune response to ICB that is present across patients regardless of response status.

We find that based on relative immune proportions in PBMC samples, responders have an increased circulating abundance of PD-1+Ki67+ CD8+ TEMRA cells post-ICB compared with non-responders^16^. Further, we noticed that many non-response proteins are expressed by macrophages in the TME, including various immunosuppressive or resistance-associated genes such as IL6^25^, CXCL8^20,21,26^ and IL10^35^, suggesting that macrophage-mediated immunosuppression is a distinguishing feature of non-response. Indeed, macrophages have been shown to contribute to immunosuppression in the melanoma TME, where M2-polarization might elicit worse outcome^44^. Furthermore, we see evidence of a T cell-macrophage interaction that may play a role in myeloid-cell plasticity that aids in the transition toward an M2-type macrophage phenotype^41,42^. Additionally, several interactions involving fibroblast signaling to cancer cells are implicated in non-response^37–40^. Our analyses suggest that fibroblast-tumor signaling axes may also be a hallmark of non-response, conferring a worse prognosis. Another subset of these genes that score high in malignant cells/melanocytes might also suggest that certain tumor-related proteins in melanoma may be involved in ICB resistance or may be unique biomarkers of tumor burden during progression.

In our plasma proteomic data, we noted proteins that are more abundant post-ICB in responders are enriched with many cytotoxicity-related proteins. Indeed, these proteins were preferentially expressed as genes in scRNA-seq data from NK cells within the tumor, emphasizing their role in cytotoxicity-related processes that are present in responding patients post-ICB. In addition to this, we noticed several epithelial proteins in this module; upon deeper analysis of their source, we inferred their likely tissue origin as the pancreas or organs of the gastrointestinal tract. The abundance of these proteins correlated with immune-related adverse events, such as pancreatitis. Pancreatitis has been noticed as an adverse event in some melanoma patients receiving ICB^45^. These data allow us to identify signatures of immunotherapy responses that correlate with specific patient toxicities.

Together, these results build a mechanistic understanding of ICB response in different compartments of the body and how signals from the circulation, tumor and healthy tissues orchestrate an immunotherapy response that distinguishes patients based on clinical outcome (**Fig. 6**).

**Figure 6.**
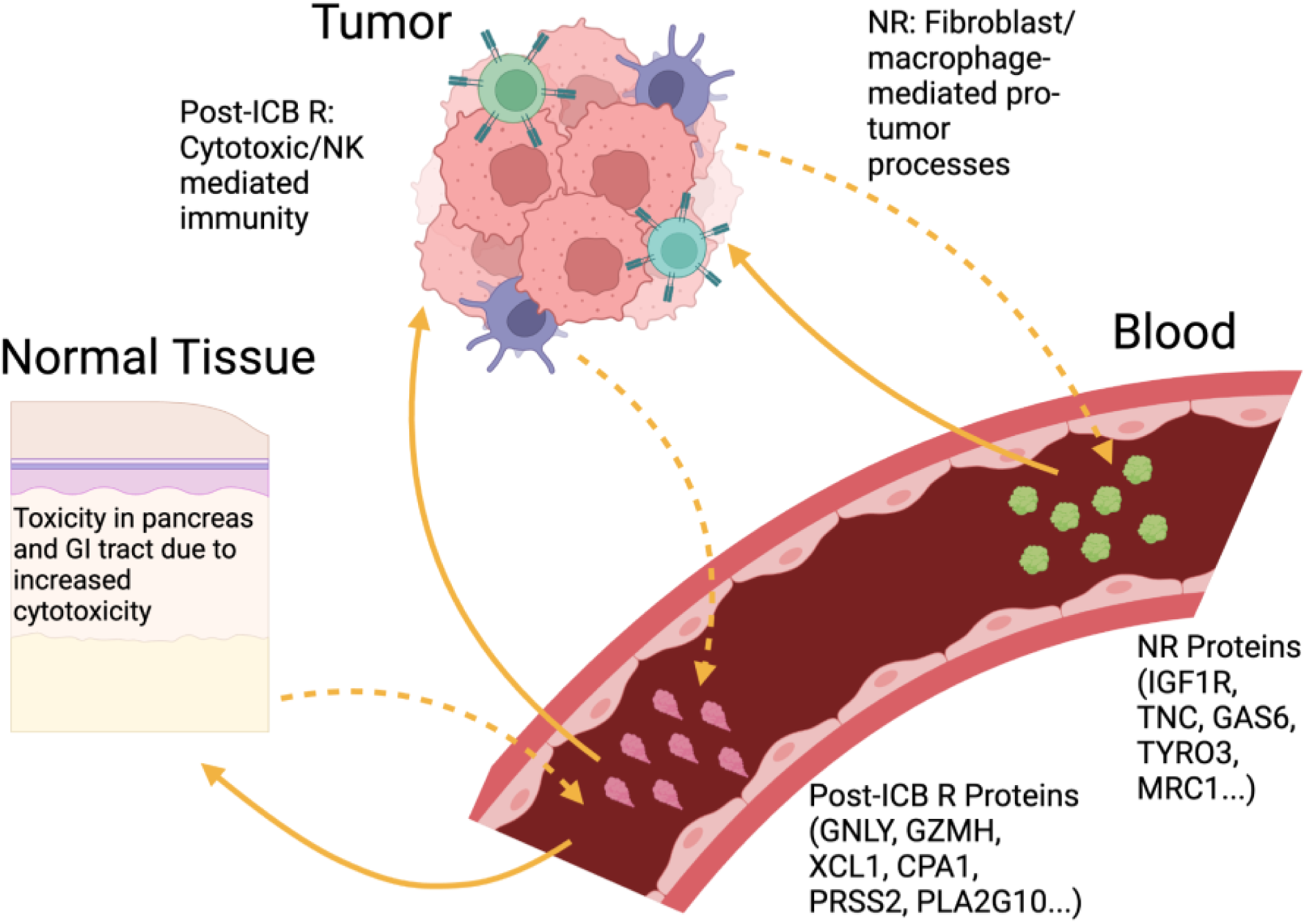
Circulating plasma proteins provide evidence for potential response and resistance processes in melanoma tumor and other normal tissues.

Our results have several limitations. We acknowledge that the proteomic panel, while comprehensive, may be biased towards specific biological processes, and that there may be other signals related to response and treatment time points that have not been revealed by this study. Further, the majority (11 out of 13) of scRNA-seq samples come from non-responding patients, potentially biasing gene expression and cell type abundance patterns. Additionally, tumor scRNA-seq samples were not time point-matched with plasma samples. Future work may involve validation of these signals using a larger sample size of scRNA-seq data derived from patient-matched tumor and PBMC samples.

## Methods

### Patient sample processing

#### Plasma, PBMC and tissue pre-processing

All patients provided written informed consent to allow the collection of blood and tumor tissue for research (Dana Farber/Harvard Cancer Center protocol identifier 11-181). Samples were collected and processed at Massachusetts General Hospital. Plasma and PBMCs were derived from 10–30cc whole blood collected in Mononuclear Cell Preparation Tubes. Plasma and PBMCs were isolated according to the BD manufacturer protocol and stored at −80C.

Tissue was collected fresh in phosphate-buffered saline. For single-cell RNA-sequencing, fresh melanoma tumors were dissociated and scRNA-seq libraries were prepared using the 10x Genomics chromium controller and reagent kits as described below. Bulk RNA sequencing was performed on tissue preserved in FFPE blocks.

Sample/patient identification numbers were not known to anyone outside of the research group.

#### Plasma protein measurements on Olink Explore

Plasma proteomics was performed in samples from 465 melanoma patient samples (248 pre-treatment, 217 on-treatment) using the Olink Explore I and II panels according to the manufacturer’s protocol. Explore combines the Proximity Extension Assay (PEA) technology with Next-generation sequencing (NGS).

In brief, the PEA technology uses matching pairs of oligonucleotide-labeled antibody probes. The PEA probes bind to target antigens producing a binding complex where the complementary oligonucleotides exist in close proximity to each other, enabling the formation of a target sequence. The dual targeting of probes has been proven to produce outstanding specificity enabling a high degree of multiplexing while maintaining sensitivity and a broad dynamic range. In the Olink Explore protocol, the target sequence is amplified in a double PCR reaction and purified before the NGS. The sequence data is processed and normalized to produce Olinks relative quantification unit Normalized Protein eXpression (NPX). The produced DNA signal functionally works as a proxy for the protein levels present in the sample.^46^

#### Peripheral blood mass cytometry

Antibodies were purchased either preconjugated to heavy-metal isotopes (Standard BioTools, formerly Fluidigm), or purified and conjugation in-house using the MaxPar X8 or MaxPar MCP9 Antibody-Labeling Kits (Standard BioTools) according to manufacturer instructions. Cryopreserved PBMC samples were thawed, washed, and resuspended in PBS supplemented with 5 mM EDTA. 5 uM Cisplatin (Sigma) was used as a live/dead stain and cells were incubated for 1 min at room temperature (RT). Cells were washed and fixed with 1.6% paraformaldehyde (PFA) in PBS supplemented with 5 mM EDTA and 0.5% BSA (PEB) for 10 min at RT. Cells were washed twice with PEB and stored at −80°C until staining.

To minimize batch-to-batch variation, samples were barcoded prior to antibody staining using a 20-Plex Pd Barcoding Kit (Standard BioTools) following manufacturer instructions. Briefly, 1 million cells were washed with MaxPar Barcode Perm Buffer (Standard BioTools) twice and barcoded with unique combinations of Pd isotopes for 30 min at room temperature on a horizontal shaker. Cells were washed twice with eBioscience permeabilization buffer (Thermo Fisher), followed by one wash with Maxpar Cell Staining Buffer (CSB, Standard BioTools) and pooled into a single tube. Antibody cocktail was prepared according to previously determined antibody titers at 25 µL of staining volume per 1 million cells. The antibody cocktail was aliquoted and stored at −80°C until staining. Before staining, Fc receptors were blocked for 10min at RT with Human Fc receptor blocking solution (Human TruStain FcX, BioLegend) followed by 60min incubation with the antibody staining cocktail at RT on a horizontal shaker. PBMCs were stained using a panel of 43 antibodies. Cells were washed twice with CSB and resuspended in intercalation solution (4% PFA in PBS and 0.5 mM iridium intercalator (Standard BioTools)) for 20 min at RT or pelleted and frozen at −80°C until acquisition.

Before acquisition, samples were washed once in CSB, twice in Cell Acquisition Solution (CAS, Standard BioTools) and filtered through a 35 µm cell strainer (Falcon). Cells were then resuspended at 0.7 million cells/mL in CAS supplemented with EQ4 element calibration beads (Standard BioTools) and acquired on a Helios mass cytometer (Standard BioTools). FCS files were processed using the R package Premessa (Parker Institute for Cancer Immunotherapy). Samples were processed in four batches and each batch contained a set of control samples. Variation between batches was normalized using CytoNorm^47^. Manual gating of FCS files was subsequently performed using CellEngine (CellCarta, Montreal, Canada) following the gating strategy as outlined in SX. Relative cell frequencies were exported as a frequency of total non-granulocytes for phenotypic immune populations (e.g. % CD-4+ T-cells / total non-granulocytes) as well as the frequency of parent for functional marker positive subsets (e.g. PD-1+ CD4+ T-cells / total CD4+ T-cells).

#### Tissue Dissociation and Library Preparation for scRNA-seq

Tumor samples were processed using the Human Tumor Dissociation Kit (Miltenyi Biotec, Cat#130-095-929) following established protocols^48,49^. Initially, tumor biopsies were minced in 1.5 mL Eppendorf tubes containing a mixture of enzymes H, R, and A (provided with the kit) and RPMI medium, and tubes were incubated in an Eppendorf thermomixer at 37°C with continuous shaking at 400 rpm for 15 minutes. Post-incubation, the dissociated cell suspension was filtered through a 50 μm filter (Sysmex, Cat#04-004-2327), followed by a wash with 5 mL of RPMI supplemented with 10% heat-inactivated fetal calf serum (FCS). The filtrate was centrifuged at 1500 rpm for 5 minutes at 4°C. After discarding the supernatant, ACK lysis buffer (Gibco, Cat#A1049201) was added to the cell pellet, gently mixed, and centrifuged under the same conditions. The ACK buffer was removed, and the cells were resuspended in RPMI with 10% heat-inactivated FCS. Cell concentration and viability were assessed using trypan blue with a Bright-Line hemocytometer (Cat#1492). For samples with viability below 70%, viable cells were enriched using a modified EasySep Dead Cell Removal (Annexin V) kit protocol (STEMCELL, Cat#17899) as described by Fang et al. (2024). After this step, cell counts and viability were reassessed, and samples were immediately prepared for single-cell RNA sequencing (scRNA-seq). Gene expression (GEX) libraries were constructed using the 10x Genomics Chromium NextGEM Single Cell V(D)J Reagents Kits v1.1 (PN-1000165, PN-1000120, PN-1000213) and the Chromium NextGEM Single Cell 5’ Reagent Kits v2 (PN-1000263, PN-1000286, PN-1000215) according to the manufacturer’s guidelines. Quality control of amplified cDNA and generated libraries included quantification using the Qubit dsDNA High Sensitivity Kit (Invitrogen, Cat#Q32854) and size distribution analysis with the Agilent High Sensitivity Bioanalyzer DNA Kit (Cat#5067-4626). Sequencing was performed on an Illumina NextSeq 500 sequencer using the 75-cycle kit (Cat#20024906). The read configurations were as follows: 1. NextGEM v1.1: Read 1 - 26 cycles; Read 2 - 55 cycles; 2. NextGEM v2: Read 1 - 26 cycles; Read 2 - 46 cycles.

### Computational analysis

#### Batch correction of Olink protein measurements performed at different times

As for one cohort in this study separate panels measuring approximately 1500 proteins each were performed at different times, bridge normalization was performed on protein measurements in this cohort to account for batch-level technical variations. Documentation for the methodology used for normalization can be found here: https://cran.r-project.org/web/packages/OlinkAnalyze/vignettes/bridging_introduction.html.

#### Statistical analysis of clinical metadata

Hypothesis tests were performed to compare clinical variables between responders and non-responders. For continuous variables, Wilcoxon rank sum test was used and for categorical variables, Fisher’s exact test was used, both in the stats package (v4.3.2) in R.

#### Survival analysis

The progression-free and overall survival or time to last follow-up along with progression or vital status, respectively, were used to fit Cox proportional hazards regression models using the survival package (v3.7.0) in R. Log-rank tests were thus used to evaluate the significance of the difference in progression-free and overall survival between responders and non-responders.

#### Linear mixed effects models

Linear mixed effects (random intercepts) models were fit for each protein using the lme4 package (v1.1.35.5) in R. All samples were used to train the model, and sample time point, patient response, and an interaction term of time point and response were used as covariates to predict protein abundance (NPX) value. Patient ID was used as a random effect. P-values for each covariate were compiled across all models, and Benjamini-Hochberg multiple hypothesis controls were used on each covariate p-value list. In separate models for each protein, covariate was added in addition to all previous covariates/random effects, and the same procedure for multiple hypothesis correction was used.

For the Teiko relative immune proportions, linear mixed effects models were used (one for each immune subset) in which sample time point, patient response and an interaction term were used as covariates to predict z-scored immune proportion. Multiple hypothesis corrected p-values are not reflected in **Table S10** as there were no p-values for any covariate that passed multiple hypothesis control.

#### Gene ontology analysis

Gene ontology analysis was performed using the gseapy package (v1.1.3) in Python, using the GO_Biological_Process_2023. The list of post-ICB proteins was used as input to generate the visualization.

#### Comparison of Ki67+PD-1+ immune cell proportions

Proportions of immune cells gated on PD-1 positivity and Ki67+ positivity were generated for each patient, and proportions were compared statistically using the ranksums function in the stats library of the scipy package (v1.10.1) in Python. Proportions were also normalized to composite scores of melanoma cell-specific genes. Composite scores were calculated using the score_genes function in the Scanpy package (v1.9.8) in Python using the NPX values of proteins in the Olink data corresponding to melanoma cell-specific genes. Melanoma cell-specific genes were determined using a workflow for Mast available on Dockstore (https://dockstore.org/workflows/github.com/MilanParikh/MAST_workflow:master?tab=info).

The significance of the melanoma cell protein score with LDH level was evaluated using the linregress function in the stats library of the scipy package (v1.10.1) in Python.

#### ScRNA-seq data preprocessing

CellRanger v7.0.0 was used to align sequencing reads to the GRCh38 human genome reference. Cell barcodes were filtered out if they met any of the following criteria: (1) fewer than 100 detected genes, (2) more than 6,000 detected genes, or (3) over 40% of unique molecular identifiers (UMIs) mapping to the mitochondrial genome.

To normalize raw counts, we applied the normalize_total function from Scanpy (v1.9.5) using a library size correction method. The top 3,000 highly variable genes were selected for principal component analysis (PCA). To address batch effects between patient samples, we employed the Harmony pipeline to generate a batch-corrected space. Major cell types were identified using Leiden clustering in Scanpy, revealing distinct populations such as B cells, CD4+ T cells, CD8+ T cells, NK cells, plasma B cells, endothelial cells, fibroblasts, macrophages, dendritic cells (DCs), cancer cells, and melanocytes.A second round of unsupervised clustering was conducted to further identify subsets within B cells, CD4+ T cells, CD8+ T cells, plasma B cells, endothelial cells, fibroblasts, macrophages, and DCs.

#### Gene scoring

For calculating composite scores across cell types, plasma samples or tissue samples in the proteomic or RNA-seq analyses, the score_genes function in the Scanpy package (v1.9.8) in Python was used.

#### Manual ligand-receptor analysis using cytokines/chemokines and their receptors

Proteins that were cytokines or chemokines, or receptors (or components of a receptor complex) for cytokines or chemokines that appeared among the linear mixed effects model proteins were compiled. CellPhoneDB’s online database was used to determine receptors or ligands for a given protein in this list (along with other members of the receptor complex if applicable). Average gene expression across major cell types for each ligand and receptor. Between each possible combination of cell type, going in both directions, the minimum of average ligand expression in one cell type and the average of receptor expression in the other cell type was determined, and interactions above a normalized expression threshold of 0.1 were considered. For receptors involving complexes, the minimum average expression of genes in the receptor complex was used to represent the average receptor expression for this calculation.

#### CellPhoneDB

The cellphonedb package (v5.0.0) in Python was used to infer cell-cell interactions. Documentation available at https://github.com/ventolab/CellphoneDB.

#### Spatial Data Acquisition and Processing

Spatial Molecular Imaging (SMI) using CosMx was performed on melanoma samples in a previous study, and we utilized publicly available data from samples #1–#4^43^ to validate our findings within a spatial context. Quality control was applied by excluding cells with fewer than 10 detected genes and genes expressed in fewer than three cells. The data were normalized to a total of 10,000 molecules per cell and log-transformed to stabilize variance. Dimensionality reduction was conducted using Principal Component Analysis (PCA), retaining the first 40 components for downstream analysis. To correct for batch effects across samples, Harmony integration^50^ (documentation available: https://github.com/immunogenomics/harmony?tab=readme-ov-file) was performed using sample identity as a covariate. Following integration, a neighborhood graph was constructed with 10 nearest neighbors, and Uniform Manifold Approximation and Projection (UMAP) was used to visualize the cellular landscape. Clustering was performed using the Leiden algorithm at a resolution of 1, based on the Harmony-corrected PCA embeddings, to identify cellular subpopulations. Harmonized clusters were annotated through differential gene expression analysis, leveraging the top 20 marker genes for each cluster. Clusters were labeled as tumor cells, T cells, B cells, fibroblasts, macrophages, keratinocytes, and endothelial cells based on marker gene expression profiles and established biological knowledge.

#### Ligand Receptor Analysis

To investigate cell-cell communication, we applied COMMOT^51^ (documentation available: https://github.com/zcang/COMMOT), a ligand-receptor interaction analysis package, to melanoma spatial transcriptomics data. The analysis was performed separately for each sample to account for sample-specific variability. We explored three categories of signaling interactions: extracellular matrix (ECM)-receptor, secreted signaling, and cell-cell contact, using the curated CellChat database for human ligand-receptor interactions. For each category, COMMOT computed spatial communication scores based on proximity (distance threshold of 75µm). Results for sender and receiver scores were extracted and saved for each sample and interaction type.

To quantify specific ligand-receptor interactions of interest, such as CXCL16-CXCR6 between macrophages and T cells, we evaluated the observed interaction frequency against a null distribution generated by permutation testing. For each sample, cell locations were mapped in two-dimensional space using their spatial coordinates. Interaction counts were determined for pairs of cells meeting the criteria of proximity (distance ≤ 75µm) and non-zero interaction scores for the specified ligand-receptor pair. To construct the null distribution, cell type labels were randomly shuffled across iterations, and interaction counts were recalculated to simulate random association.

The statistical significance of observed interactions was assessed by comparing the observed interaction count to the null distribution. A p-value was computed as the proportion of null interaction counts greater than or equal to the observed count.

#### Substitute genes for TNC and TYRO that were present in the CosMx panel

TNC and TYRO were not available in the gene panel of the CosMx data used and as such we needed to find genes that were expressed uniquely in TNC+ fibroblasts and TYRO+ melanoma cells. To find a substitute gene for TNC, we subsetted the data to fibroblasts and performed differential gene expression analysis comparing TNC+ fibroblasts (normalized TNC expression > 0) and TNC-fibroblasts using the score_genes function in the Scanpy package (v1.9.8) in Python. Top differentially expressed genes for TNC+ fibroblasts were examined, and THBS1 was chosen because it was highly expressed in TNC+ fibroblasts and had almost absent expression in TNC-fibroblasts, in addition to being present in the CosMx panel. The same procedure was repeated for TYRO3+ melanoma cells, and NRP2 was chosen as a substitute this way.

#### GTEx analysis

Bulk RNA-seq data was downloaded from the GTEx portal (https://gtexportal.org/home/). Q normalization was used across samples using the qnorm package (v0.8.1) in Python.

#### Toxicity Annotations

Patients of reviewed cases were classified by internists and medical oncologists as having ICI-related pancreatitis based on documentation by treating clinicians, laboratory findings, and/or radiology reports.

#### Statistical analysis of toxicity annotations against Olink protein abundances

Statistical significance of the association between the presence of a particular toxicity annotation and proteomic abundance was determined using the ranksums function in the stats library of the scipy package (v1.10.1) in Python. Each epithelial protein from the post-ICB protein module favored in responders was considered. Benjamini-Hochberg multiple hypothesis correction was performed on the list of resulting p-values separately for each toxicity annotation. Adjusted p-values below 0.1 were considered statistically significant.

## Data Availability

All data produced in the present work are contained in the manuscript

## Acknowledgments

We would like to thank funding support from the Prostate Cancer Foundation Young Investigator Award (RJP), Dana Farber Cancer Institute / Harvard CancerCare GI SPORE Career Enhancement Award (AM), Sky Foundation Pancreatic Cancer Research Grant (AM), Doris Duke Charitable Foundation Physician Scientist Fellowship (AM) and DF/HCC K12 (K12CA087723) Paul Calabresi Award for Clinical Oncology (AM). We acknowledge funding provided by the Massachusetts Life Sciences Center Research Infrastructure Program in support of the Mass General Cancer Center Tumor Cartography Center and the Dr. Miriam and Sheldon G. Adelson Medical Research Foundation.

## Supplemental Figure Legends

**Figure S1. a,** Breakdown of treatment type received by each patient whose plasma underwent proteomic profiling. **b,** Number of patients in the melanoma immunotherapy cohort for which Olink (plasma proteomics), Teiko (PBMC relative immune cell proportions), Bulk RNA-seq and single-cell RNA-seq were available. Numbers in overlapping segments represent quantities of patients for which more than one data modality was available. **c,** Number of patients from the plasma proteomic cohort for which one or both of pre-treatment baseline (BL) and 6 weeks on-treatment (6W) time points were available. **d,** Number of patients from the PBMC relative immune proportions (Teiko) cohort for which one or both of pre-treatment baseline (BL) and 6 weeks on-treatment (6W) time points were available.

**Figure S2. a,** Distribution of normalized protein expression (NPX) means across samples. **b,** Coefficient of variation (CoV) for each protein plotted against its mean.

**Figure S3.** Overlap of proteins with significant response or time point effects in original linear mixed effects models vs. linear mixed effects models with an additional subtype covariate. Top left, overlapping post-ICB proteins; bottom left, overlapping pre-ICB proteins; top right, overlapping non-responder proteins; bottom right, overlapping responder proteins.

**Figure S4. a,** Comparison of response coefficients in original and cutaneous-only linear mixed effects models, among proteins from original linear mixed effects models that had significant response effects. **b,** Comparison of responder distribution skewness in total and cutaneous-only sample groups among proteins from original linear mixed effects models that had significant response effects. **c,** Comparison of non-responder distribution skewness in total and cutaneous-only sample groups among proteins from original linear mixed effects models that had significant response effects.

**Figure S5. a,** Comparison of response coefficients in original and SD responder linear mixed effects models, among proteins from original linear mixed effects models that had significant response effects. **b,** Comparison of responder distribution skewness in total and SD-responder sample groups among proteins from original linear mixed effects models that had significant response effects. **c,** Comparison of non-responder distribution skewness in total and SD-responder sample groups among proteins from original linear mixed effects models that had significant response effects.

**Figure S6.** Top gene ontology (GO) enrichment terms on the on-treatment proteins using the GO Biological Processes 2023 library. GO score shown in x-axis, percent of on-treatment genes in the gene set shown by the size of each dot, −log(FDR) of enrichment shown by color of each dot.

**Figure S7. a,** Hierarchically-clustered heatmap of cosine similarity between composite protein group scores from linear mixed effects models (and hierarchical clustering of on-treatment proteins) and PBMC relative immune proportions. **b,** PBMC relative immune proportions linear mixed effects model coefficients, ordered by magnitude and sign, with −log(p-value) shown by color of dot. Left, response effects. Middle, time point effects. Right, interaction effects. **c,** Correlation of LDH level and composite malignant proteomic score across samples. **d,** Association of KI67+PD1+ CD8 TEMRA cells with treatment response on-treatment, normalized by LDH level. Wilcoxon rank-sum test, p = 0.046. **e,** Association of overall KI67+PD1+ CD8 T cells with treatment response on-treatment, normalized by LDH level. Wilcoxon rank-sum test, p = 0.181.

**Figure S8. a,** Hierarchically-clustered heatmap showing presence of non-response proteins in each disease cluster (Deng et al., 2024). Colored red if protein is present in the cluster. **b,** Number of non-responder proteins present in each disease cluster (Deng et al., 2024).

**Figure S9.** Cell type composition in each single-cell RNA-seq sample.

**Figure S10. a,** Heatmap visualization of expression of receptors for cytokines, chemokines and other secreted immune-related genes across single-cell subtypes. **b,** Flow diagram showing potential interactions involving cytokines and chemokines among non-responder proteins based on minimum expression of only receptors in receiver cells.

**Figure S11.** CellPhoneDB output plot showing interaction score (shown by color), −log(p-value) (shown by size) and significance (whether the dot is circled in red) across cell type pairs for interactions involving post-ICB proteins.

**Figure S12.** CellPhoneDB output plot showing interaction score (shown by color), −log(p-value) (shown by size) and significance (whether the dot is circled in red) across cell type pairs for interactions involving pre-ICB proteins (top), non-responder proteins (middle), and non-responder, post-ICB proteins (bottom).

## Supplemental Table Legends

**Table S1.** Clinical metadata of patient samples across plasma proteomics cohort.

**Table S2.** Plasma proteomics normalized proteomic expression (NPX) matrix across patient samples.

**Table S3.** Relative immune proportions profiled from PBMC samples across patient samples.

**Table S4.** Proteins that had at least one significant linear mixed effect model covariate. A one is indicated in the appropriate column if that protein was significant for a particular term. Slope coefficients are shown for each term in the rightmost columns.

**Table S5.** Proteins that had at least one significant covariate in the linear mixed effects model with the added subtype term. A one is indicated in the appropriate column if that protein was significant for a particular term. Slope coefficients are shown for each term in the rightmost columns.

**Table S6.** Comparison of proteins significant for response in the original linear mixed effects models and in the linear mixed effects models with the added subtype term.

**Table S7.** Proteins that had at least one significant covariate in the cutaneous-only linear mixed effects models. P-values are indicated in the appropriate column for each covariate. Slope coefficients are shown for each term in the rightmost columns.

**Table S8.** Proteins that had at least one significant covariate in the SD responder linear mixed effects models. P-values are indicated in the appropriate column for each covariate. Slope coefficients are shown for each term in the rightmost columns.

**Table S9.** Top 30 genes in each post-ICB protein non-negative factorization module with six components.

**Table S10.** P-values and slope coefficients for linear mixed effects models predicting PBMC relative immune proportions.

**Table S11.** Non-responder proteins that overlapped with each Deng et al., 2024 module.

**Table S12.** Diseases in each Deng et al., 2024 module, ordered by modules with most overlapping non-responder proteins.

**Table S13.** CellPhoneDB output file showing cell-cell interactions with significant means.

