## Supplemental Figures for "Multimodal blood based profiling reveals insights into mechanisms of immunotherapy resistance"

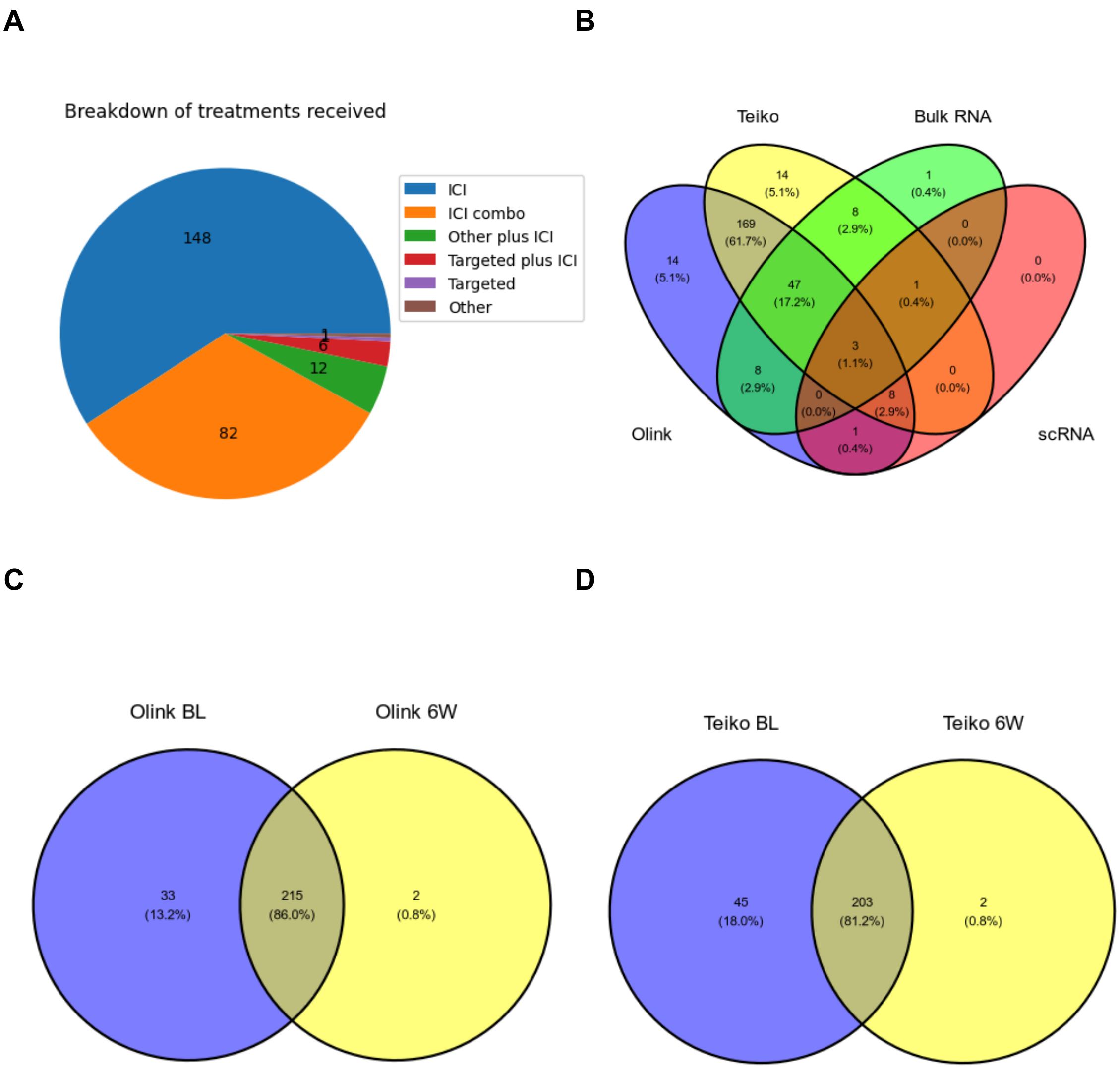
A
Breakdown of treatments received
148
82
12
6
1
ICI
ICI combo
Other plus ICI
Targeted plus ICI
Targeted
Other
B
Teiko
Bulk RNA
Olink
scRNA
14 (5.1%)
1 (0.4%)
169 (61.7%)
8 (2.9%)
0 (0.0%)
14 (5.1%)
47 (17.2%)
1 (0.4%)
0 (0.0%)
3 (1.1%)
8 (2.9%)
0 (0.0%)
0 (0.0%)
8 (2.9%)
1 (0.4%)
C
Olink BL
Olink 6W
33 (13.2%)
215 (86.0%)
2 (0.8%)
D
Teiko BL
Teiko 6W
45 (18.0%)
203 (81.2%)
2 (0.8%)

### Figure S1

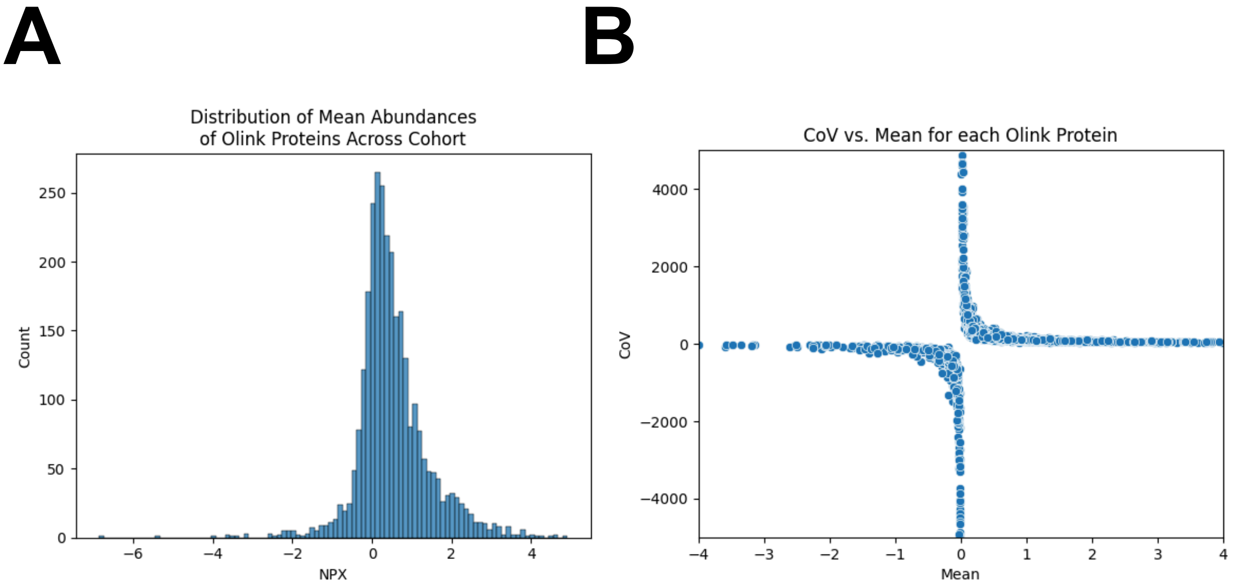
A
B
Distribution of Mean Abundances
of Olink Proteins Across Cohort
Count
NPX
CoV vs. Mean for each Olink Protein
CoV
Mean

Figure S2

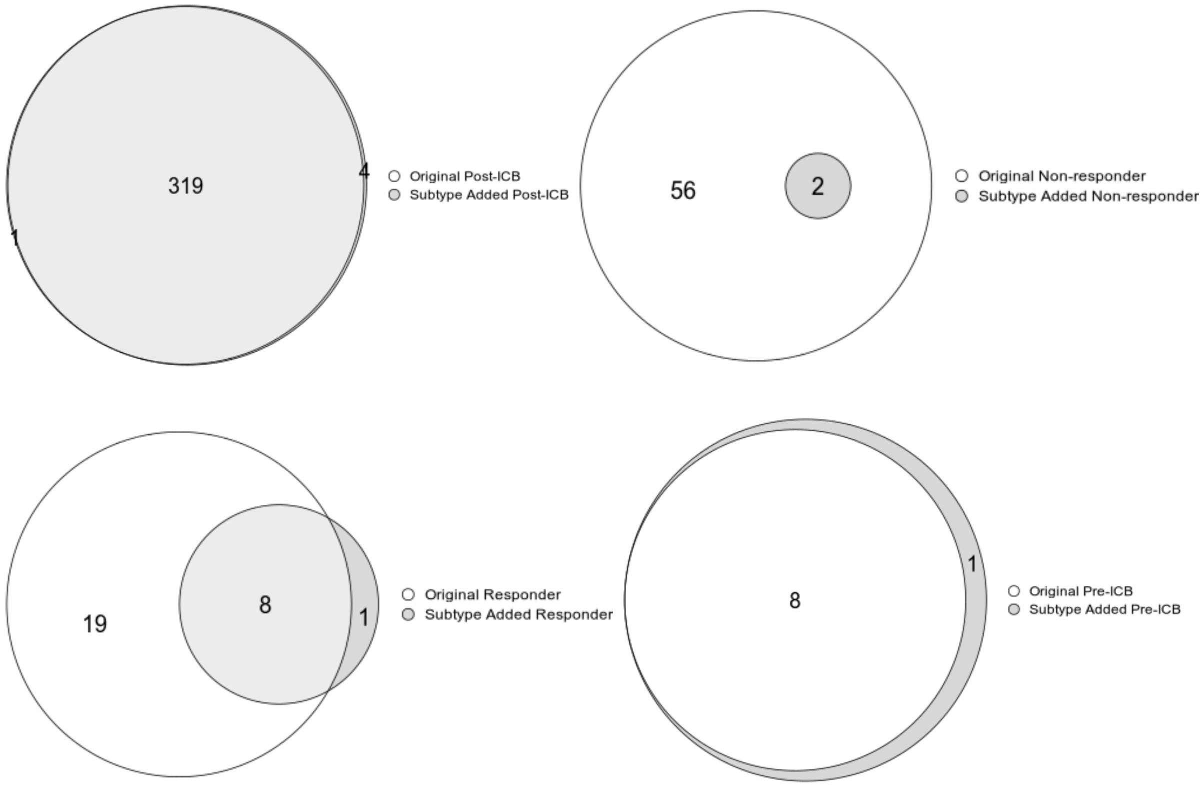
319
4
1
Original Post-ICB
Subtype Added Post-ICB
56
2
Original Non-responder
Subtype Added Non-responder
19
8
1
Original Responder
Subtype Added Responder
8
1
Original Pre-ICB
Subtype Added Pre-ICB

### Figure S3

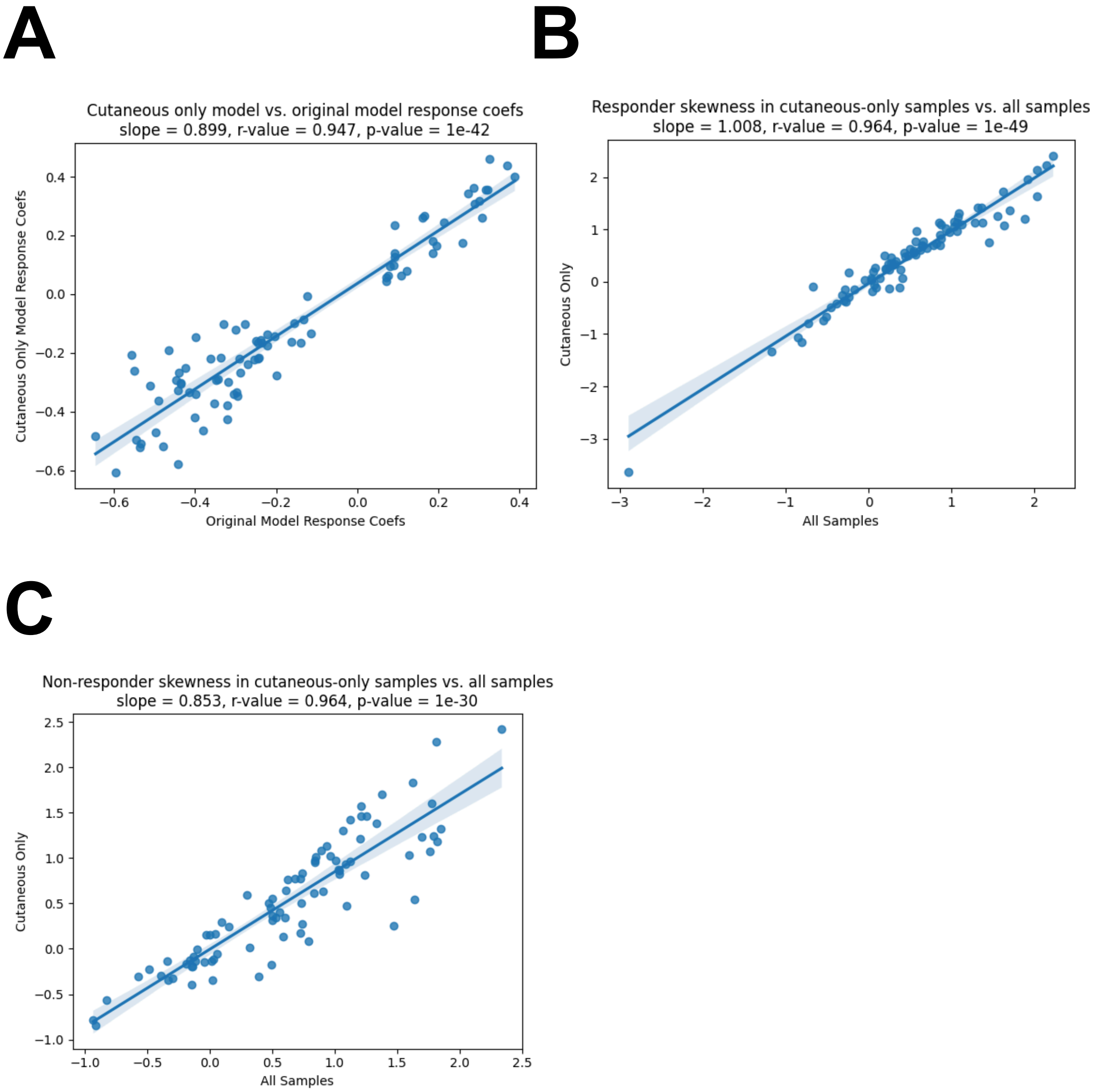
A
Cutaneous only model vs. original model response coefs
slope = 0.899, r-value = 0.947, p-value = 1e-42
Cutaneous Only Model Response Coefs
Original Model Response Coefs
B
Responder skewness in cutaneous-only samples vs. all samples
slope = 1.008, r-value = 0.964, p-value = 1e-49
Cutaneous Only
All Samples
C
Non-responder skewness in cutaneous-only samples vs. all samples
slope = 0.853, r-value = 0.964, p-value = 1e-30
Cutaneous Only
All Samples

**Figure S4**

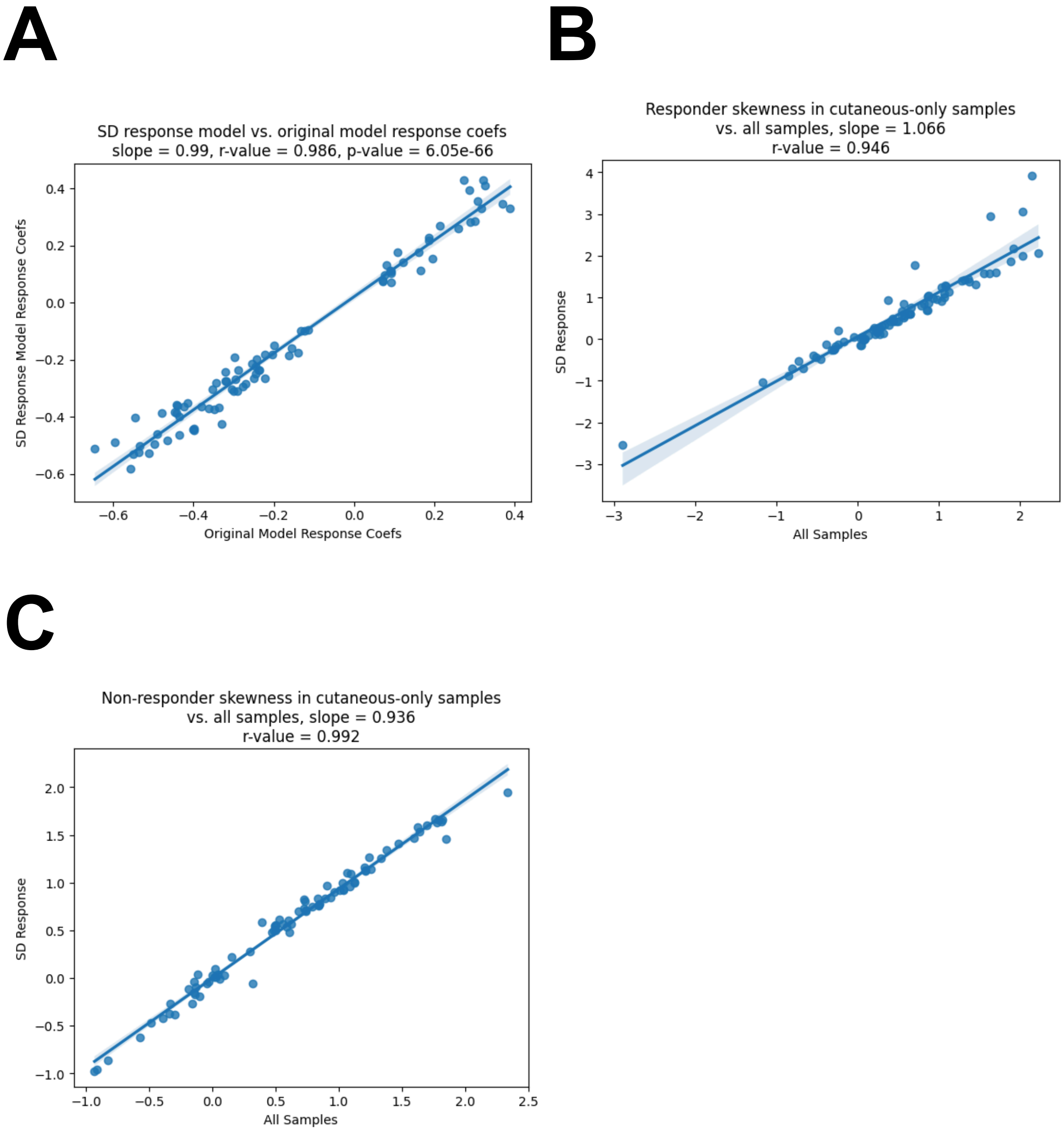
A
SD response model vs. original model response coefs
slope = 0.99, r-value = 0.986, p-value = 6.05e-66
SD Response Model Response Coefs
Original Model Response Coefs
B
Responder skewness in cutaneous-only samples
vs. all samples, slope = 1.066
r-value = 0.946
SD Response
All Samples
C
Non-responder skewness in cutaneous-only samples
vs. all samples, slope = 0.936
r-value = 0.992
SD Response
All Samples

Figure S5

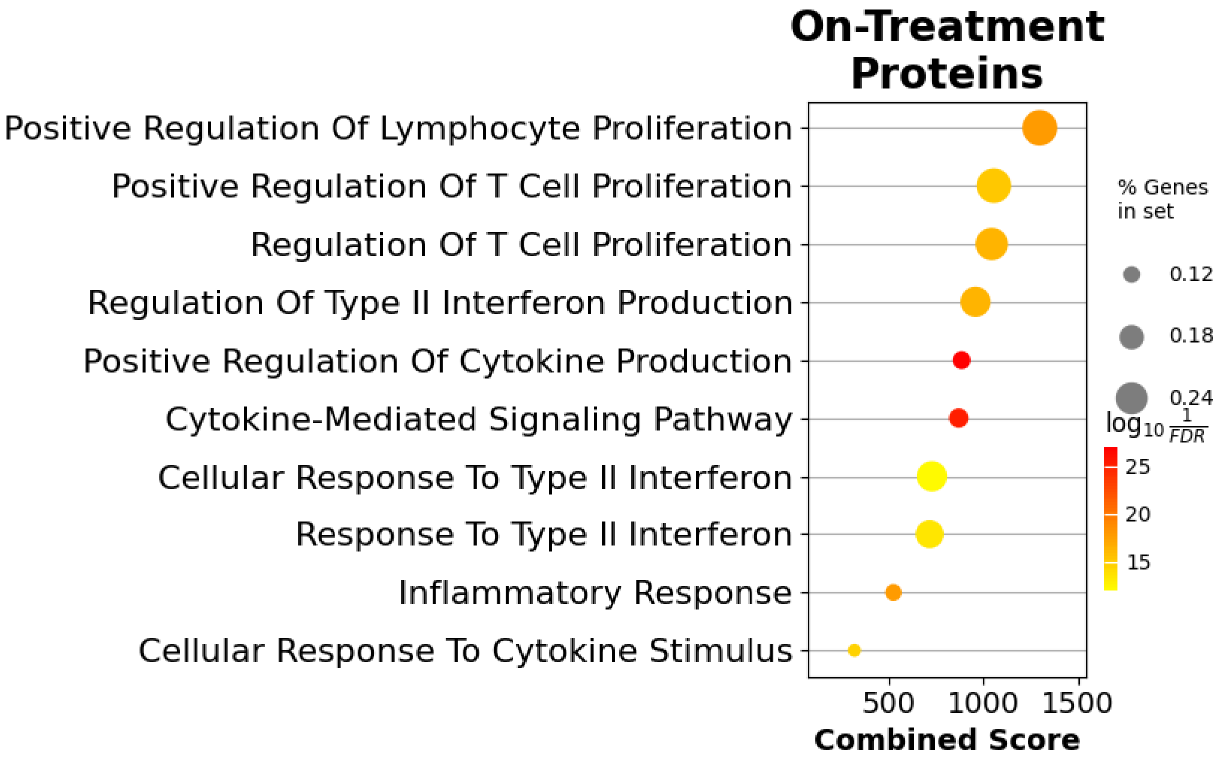
On-Treatment Proteins
Positive Regulation Of Lymphocyte Proliferation
Positive Regulation Of T Cell Proliferation
Regulation Of T Cell Proliferation
Regulation Of Type II Interferon Production
Positive Regulation Of Cytokine Production
Cytokine-Mediated Signaling Pathway
Cellular Response To Type II Interferon
Response To Type II Interferon
Inflammatory Response
Cellular Response To Cytokine Stimulus
500
1000
1500
Combined Score
% Genes in set
0.12
0.18
0.24
$\log_{10} \frac{1}{FDR}$
25
20
15

### Figure S6

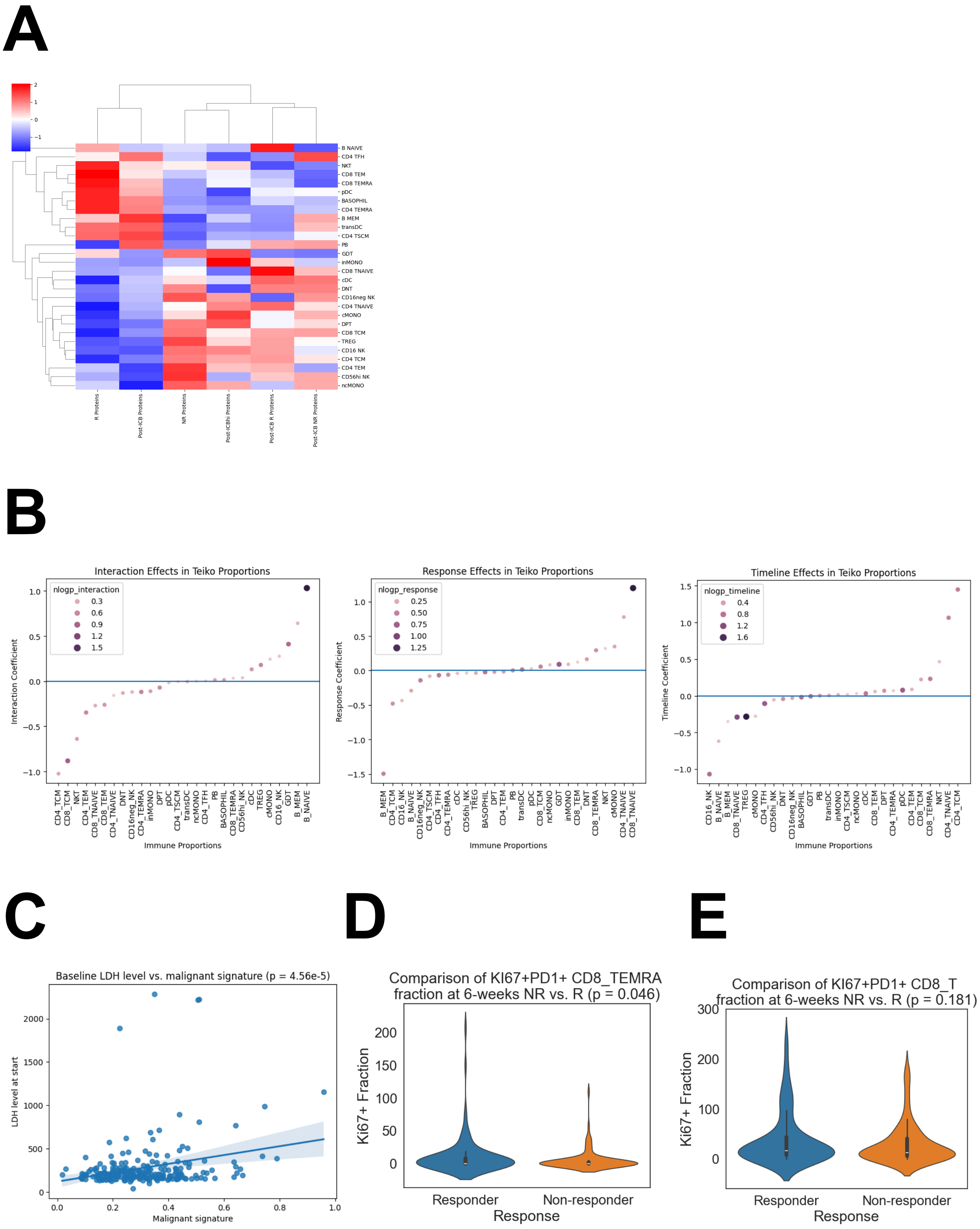
A
R Proteins
Post-ICB R Proteins
NR Proteins
Post-ICBhi Proteins
Post-ICB R Proteins
Post-ICB NR Proteins
B
Interaction Effects in Teiko Proportions
Interaction Coefficient
Immune Proportions
nlogp_interaction
Response Effects in Teiko Proportions
Response Coefficient
nlogp_response
Timeline Effects in Teiko Proportions
Timeline Coefficient
nlogp_timeline
C
Baseline LDH level vs. malignant signature (p = 4.56e-5)
LDH level at start
Malignant signature
D
Comparison of KI67+PD1+ CD8_TEMRA fraction at 6-weeks NR vs. R (p = 0.046)
Ki67+ Fraction
Responder
Non-responder
Response
E
Comparison of KI67+PD1+ CD8_T fraction at 6-weeks NR vs. R (p = 0.181)
Ki67+ Fraction
Responder
Non-responder
Response

### Figure S7

A

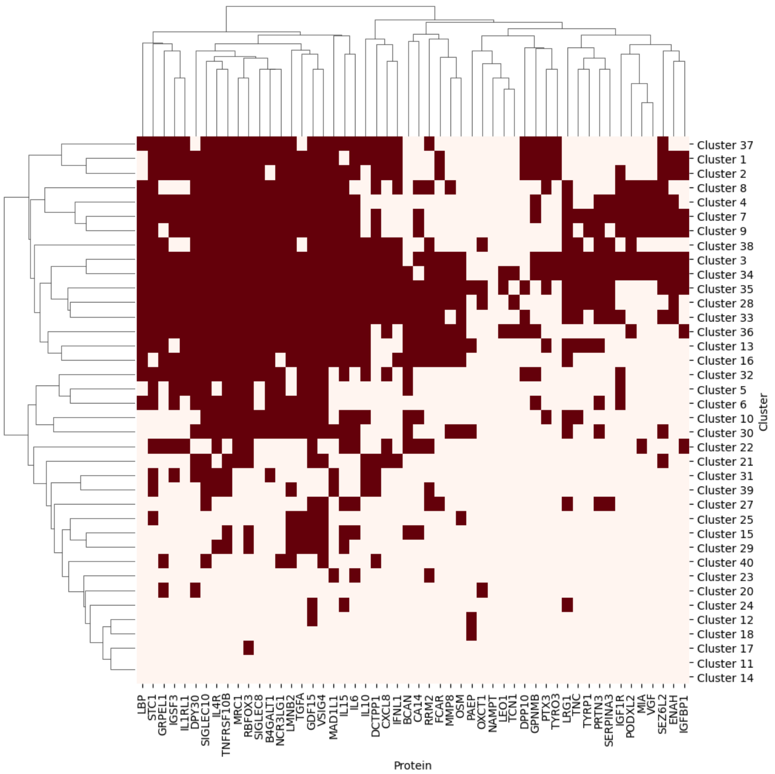
Cluster 37
Cluster 1
Cluster 2
Cluster 8
Cluster 4
Cluster 7
Cluster 9
Cluster 38
Cluster 3
Cluster 34
Cluster 35
Cluster 28
Cluster 33
Cluster 36
Cluster 13
Cluster 16
Cluster 32
Cluster 5
Cluster 6
Cluster 10
Cluster 30
Cluster 22
Cluster 21
Cluster 31
Cluster 39
Cluster 27
Cluster 25
Cluster 15
Cluster 29
Cluster 40
Cluster 23
Cluster 20
Cluster 24
Cluster 12
Cluster 18
Cluster 17
Cluster 11
Cluster 14
Cluster
LBP
STC1
GRPEL1
IGSF3
IL1RL1
DPY30
SIGLEC10
IL4R
TNFRSF10B
MRC1
RBFOX3
SIGLEC8
B4GALT1
NCR3LG1
LMNB2
TGFA
GDF15
VSIG4
MAD1L1
IL15
IL6
IL10
DCTPP1
CXCL8
IFNL1
BCAN
CA14
RRM2
FCAR
MMP8
OSM
PAEP
OXCT1
NAMPT
LEO1
TCN1
DPP10
GPNMB
PTX3
TYRO3
LRG1
TNC
TYRP1
PRTN3
SERPINA3
IGF1R
PODXL2
MIA
VGF
SEZ6L2
ENAH
IGFBP1
Protein

B

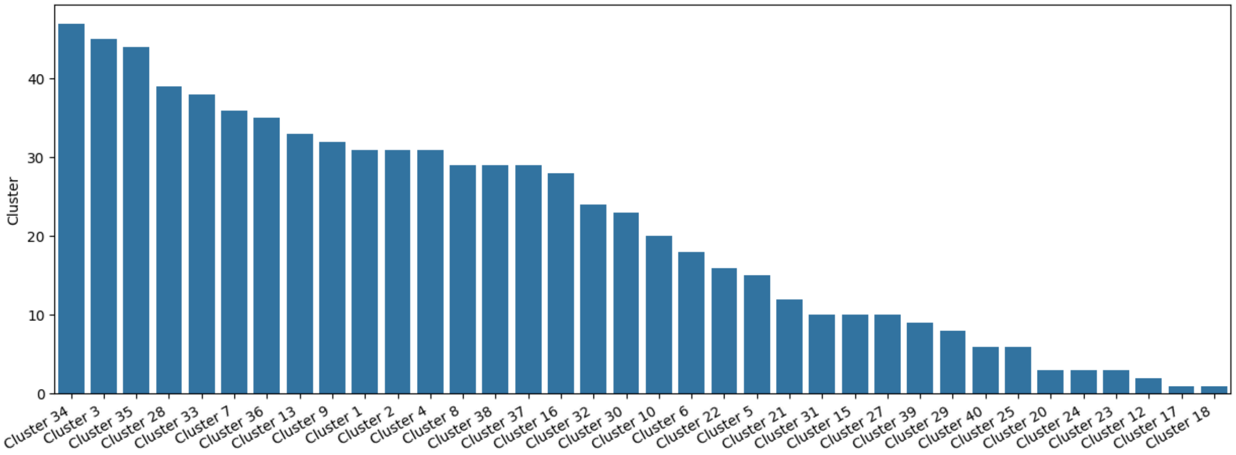
Cluster
0
10
20
30
40
Cluster 34
Cluster 3
Cluster 35
Cluster 28
Cluster 33
Cluster 7
Cluster 36
Cluster 13
Cluster 9
Cluster 1
Cluster 2
Cluster 4
Cluster 8
Cluster 38
Cluster 37
Cluster 16
Cluster 32
Cluster 30
Cluster 10
Cluster 6
Cluster 22
Cluster 5
Cluster 21
Cluster 31
Cluster 15
Cluster 27
Cluster 39
Cluster 29
Cluster 40
Cluster 25
Cluster 20
Cluster 24
Cluster 23
Cluster 12
Cluster 17
Cluster 18

### Figure S8

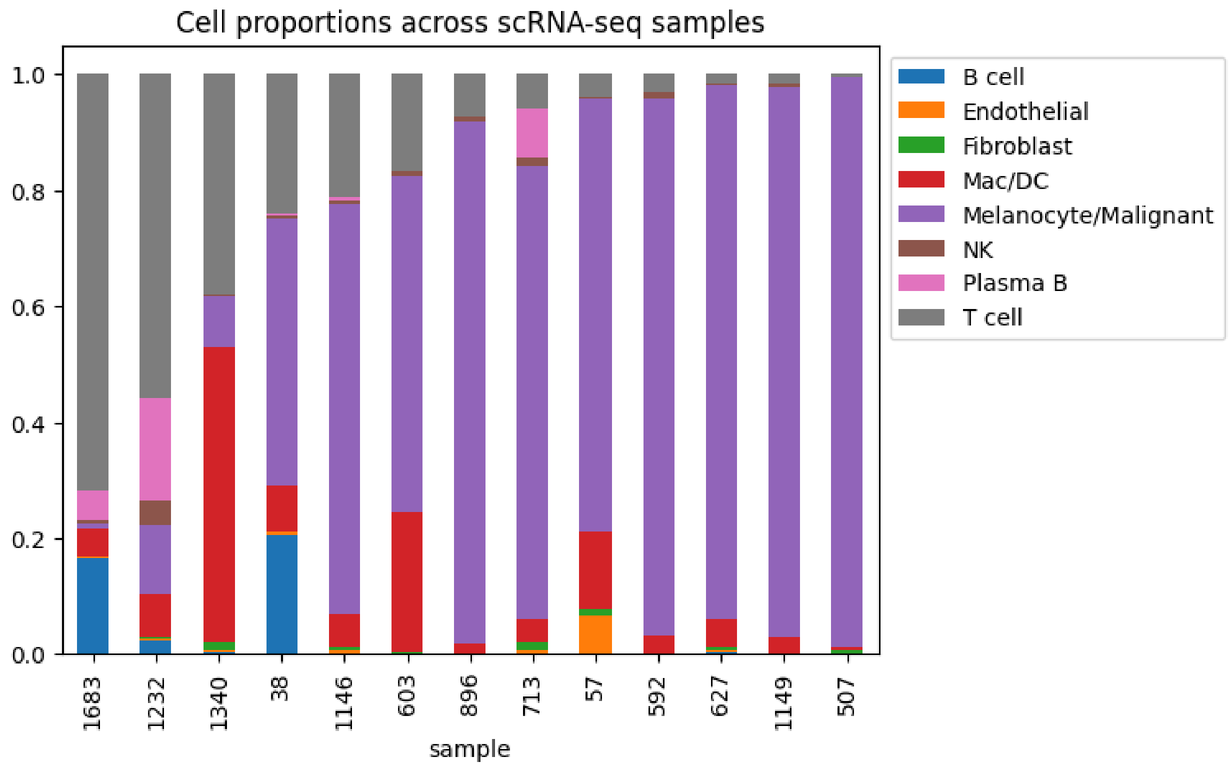
Cell proportions across scRNA-seq samples
1.0
0.8
0.6
0.4
0.2
0.0
1683
1232
1340
38
1146
603
896
713
57
592
627
1149
507
sample
B cell
Endothelial
Fibroblast
Mac/DC
Melanocyte/Malignant
NK
Plasma B
T cell

### Figure S9

A

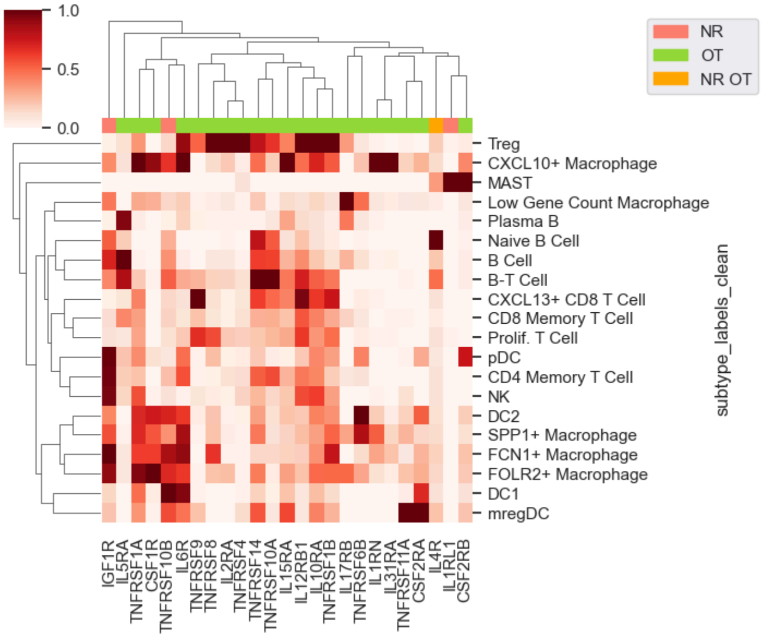


B

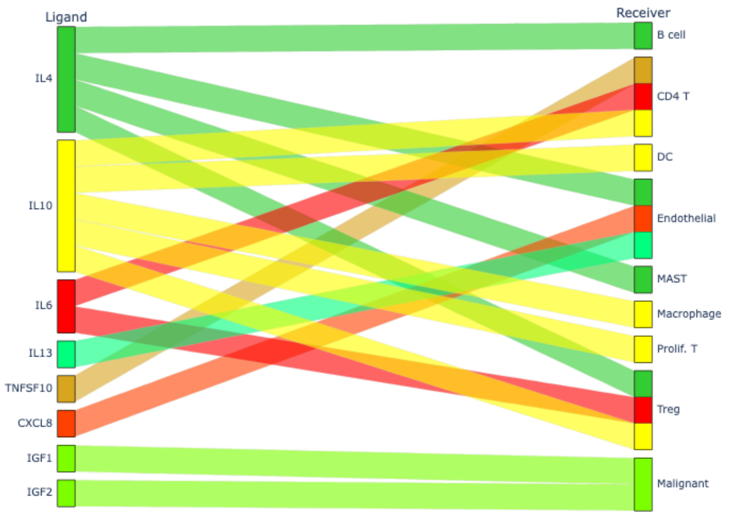


### Figure S10

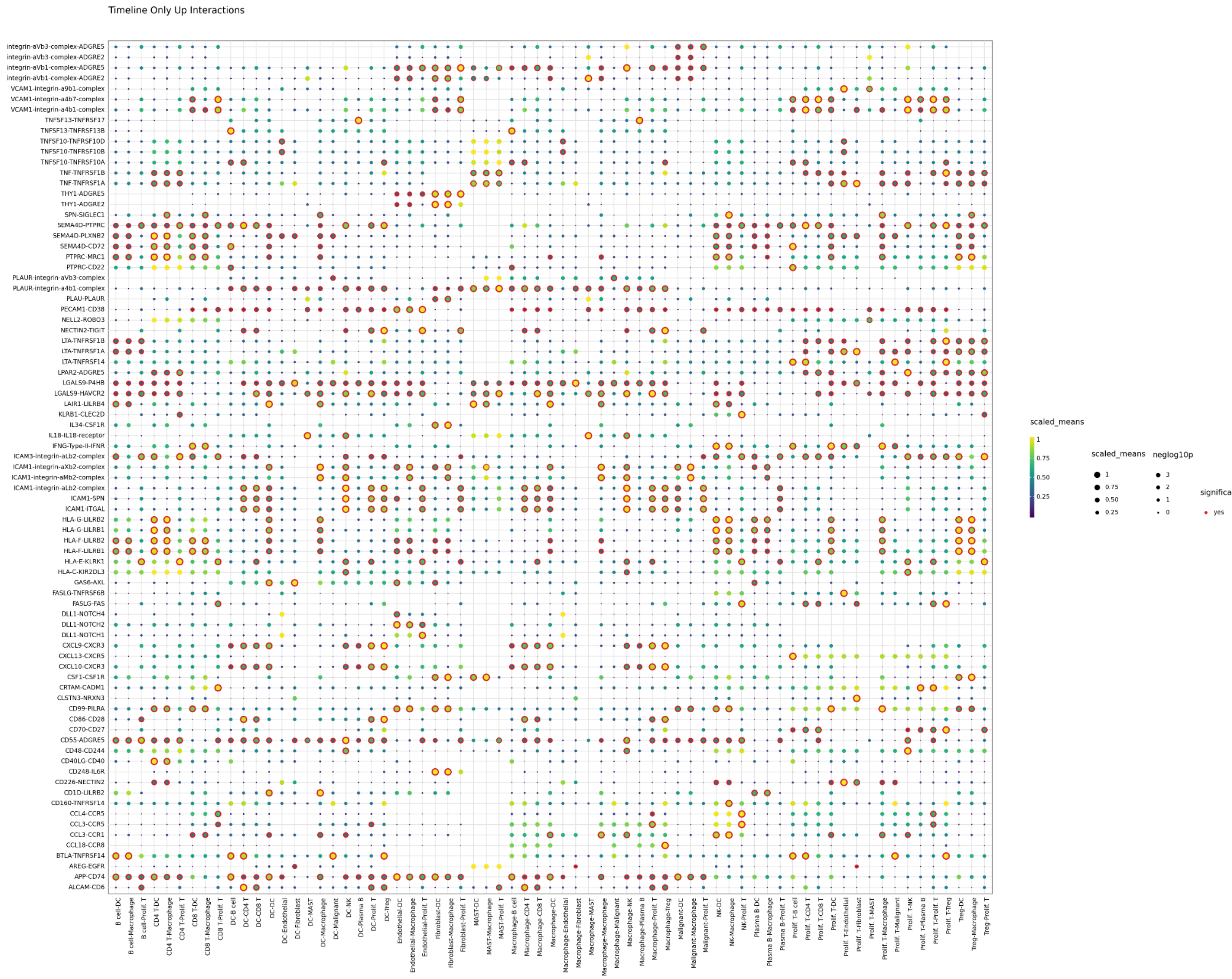

### Figure S11

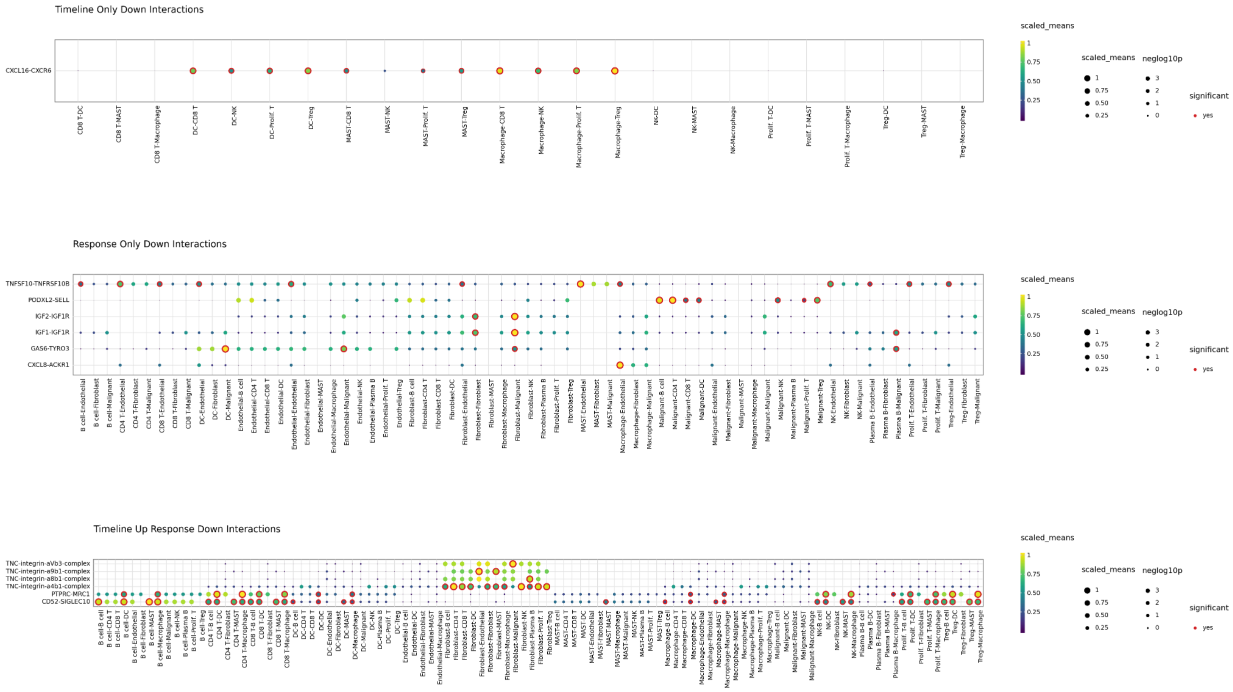
Timeline Only Down Interactions
CXCL16-CXCR6
CD8 T-DC
CD8 T-MAST
CD8 T-Macrophage
DC-CD8 T
DC-NK
DC-Prolif. T
DC-Treg
MAST-CD8 T
MAST-NK
MAST-Prolif. T
MAST-Treg
Macrophage-CD8 T
Macrophage-NK
Macrophage-Prolif. T
Macrophage-Treg
NK-DC
NK-MAST
NK-Macrophage
Prolif. T-DC
Prolif. T-MAST
Prolif. T-Macrophage
Treg-DC
Treg-MAST
Treg-Macrophage
scaled_means
neglog10p
significant
yes
Response Only Down Interactions
TNFSF10-TNFRSF10B
PODXL2-SELL
IGF2-IGF1R
IGF1-IGF1R
GAS6-TYRO3
CXCL8-ACKR1
Timeline Up Response Down Interactions
TNC-integrin-aVb3-complex
TNC-integrin-a9b1-complex
TNC-integrin-a8b1-complex
TNC-integrin-a4b1-complex
PTPRC-MRC1
CD52-SIGLEC10

### Figure S12
